# Transcranial Pulse Stimulation Enhances Dexterity in Parkinson’s Disease: A Randomized Sham-Controlled Clinical Trial

**DOI:** 10.1101/2025.07.07.25330807

**Authors:** Eva Matt, Nina Plischek, Michael Mitterwallner, Sonja Radjenovic, Alexandra Weber, Alina Domitner, Gregor Dörl, Anna Zettl, Sarah Osou, Anna Leitgeb, Agnes Santer, Viktoria Winkler, Julia Mandelburger, Heidemarie Zach, Roland Beisteiner

**Affiliations:** Functional Brain Diagnostics and Therapy, Department of Neurology, Medical University of Vienna, Vienna, Austria; Department of Neurology, Medical University of Vienna, Vienna, Austria

**Author notes:** Corresponding author: Roland Beisteiner.

## Abstract

**Background:** Transcranial focused ultrasound, capable of reaching deep brain areas with exceptional precision, represents cutting-edge technology in non-invasive brain stimulation. Current understanding of the neuromodulatory effects of focused ultrasound in Parkinson’s disease (PD) is based on small-scale, exploratory studies, with no available data on functional or structural magnetic resonance imaging (MRI) outcomes.

**Methods:** In the largest prospective, randomized, sham-controlled clinical trial to date on focused ultrasound neuromodulation in PD, the efficacy of transcranial pulse stimulation (TPS) in alleviating sensorimotor symptoms in patients with mild PD was evaluated. In a crossover design, 30 participants underwent six sessions of both verum and sham TPS targeting the sensorimotor network. Clinical and neuropsychological assessments were administered at baseline, one day post stimulation, and one month post stimulation for both conditions. Structural and functional brain changes were assessed using task-based functional MRI and diffusion tensor imaging (DTI).

**Results:** Manual dexterity, measured via the coin rotation task, showed significantly larger improvement following verum compared to sham TPS. In contrast, gross motor symptoms improved similarly after both verum and sham TPS. Functional MRI during a coin rotation task revealed increased activation in the sensorimotor network following verum stimulation compared to sham. DTI indicated enhanced structural integrity in the primary somatosensory white matter following verum TPS compared to sham.

**Conclusion:** These findings indicate that TPS produces behavioral, neurophysiological, and structural effects within the targeted sensorimotor network, and may represent a safe and effective adjunct to standard PD therapies.

**Trial registration:** This trial is registered on ClinicalTrials.gov (NCT04333511).

## Introduction

Parkinson’s disease (PD) is a progressive neurodegenerative movement disorder that is clinically diagnosed based on hallmark motor symptoms, including bradykinesia, resting tremor, rigidity, and postural instability [1]. Neuropathologically, PD is characterized by the accumulation of Lewy pathology, first appearing in brain stem nuclei during prodromal stages and spreading to limbic and cortical regions as the disease advances [2,3]. Motor symptoms are caused by the progressive degeneration of the nigrostriatal pathway, inducing widespread functional and structural alterations in subcortical and cortical motor networks [4].

Levodopa remains the gold standard for treating motor symptoms in PD, alleviating particularly bradykinesia and rigidity for several hours (*on* time) before motor symptoms return (*off* time). Over time, these motor fluctuations become more pronounced, necessitating higher doses, which are frequently associated with increased side effects [3]. Dopamine agonists represent an alternative treatment option, although dose-dependent side effects are also common. Deep brain stimulation (DBS), which delivers electric current to the subthalamic nucleus or globus pallidus via implanted intracranial leads, significantly increases *on* time and often permits a reduction in dopaminergic medication dosage. Recently, incisionless ablation of the subthalamic nucleus or the globus pallidus using high-intensity focused ultrasound demonstrated improvements in motor function and reductions in dyskinesia in PD [5,6]. Nonetheless, these procedures have been associated with adverse events, including dyskinesia, speech disturbances, and gait impairments [3].

Non-invasive brain stimulation (NIBS) presents a promising alternative for the treatment of PD, offering a favorable safety profile with only mild side effects and the potential for persistent effects through modulation of neuroplasticity [4,7]. Randomized controlled trials investigating repetitive transcranial magnetic stimulation (rTMS) and transcranial direct current stimulation (tDCS) have reported improved motor symptoms, as well as alleviated dyskinesia and depression (for review, see [4]). However, even successful rTMS and tDCS protocols are not as effective as levodopa and long-term benefits remain rather modest. Moreover, these NIBS approaches are limited by coarse targeting and are restricted to cortical regions. Transcranial application of ultrasound permits access to deep brain structures, such as the substantia nigra or the striatum. These areas were sonicated using unfocused low-intensity pulsed ultrasound in a sham-controlled trial in 56 PD patients with cognitive impairment, reporting improvements of motor symptoms, cognitive abilities, and mood following 40 stimulation sessions [8].

In contrast, low-intensity MRI-guided *focused* ultrasound, with its capability for precise targeting of cortical and deep brain structures, represents the cutting edge of clinical neuromodulation [9,10]. In PD, current investigations utilizing this technology include only exploratory, small-scaled trials involving 10 to 20 patients. They studied the effects of transcranial focused ultrasound (FUS) [11–13] and transcranial pulse stimulation (TPS) [14,15]. Targeting the primary motor cortex (M1), amplitudes of motor evoked potentials (MEPs) were increased following repeated applications of theta burst FUS (tbFUS, 5 Hz pulse repetition) in PD patients *off* dopaminergic therapy [11] and after a single application in PD patients *on* [12]. Sonication of the globus pallidus with tbFUS and 10 Hz FUS increased power of local field potentials, measured via DBS leads, implying target-specific upregulation of the motor network excitability via FUS [13]. Immediately after a single TPS session targeting the M1, resting tremor amplitude was significantly reduced as compared to sham [15]. However, this difference was not significant 24 h after the stimulation, suggesting that the benefit of a single TPS session relative to placebo is limited in time. Improvements in global motor scores, such as the Unified Parkinson’s disease Rating Scale (UPDRS) or its revision Movement Disorder Society (MDS)-UPDRS, were observed in a retrospective open-label study of TPS in 20 patients with PD [14], but comparisons to sham did not reach statistical significance in other small, explorative trials [11,15].

Since larger scale studies are important for judgement of clinical effects of focused ultrasound, we here present the currently largest prospective, randomized, sham-controlled, crossover study involving 30 patients with PD (**Figure 1A**). We investigated the effects of six sessions of TPS targeting the bilateral sensorimotor network (**Figure 1B, C**). Primary outcomes included changes in global motor symptoms, assessed using the MDS-UPDRS-III, and manual dexterity, measured via a coin rotation task. Given that the effects of ultrasound neuromodulation on functional and structural magnetic resonance imaging (MRI) measures in PD remain unclear, we conducted comprehensive MRI assessments, including task-based functional MRI (fMRI) during a simple motor task (finger tapping) and a complex sensorimotor task (coin rotation), as well as diffusion tensor imaging (DTI) to evaluate changes in structural integrity of the white matter tracts. Motor evaluations were complemented by neuropsychological assessments probing global cognition, depression, and activities of daily living, with assessments at baseline, post stimulation, and one-month post-stimulation to investigate long-term clinical efficacy.

**Figure 1.**
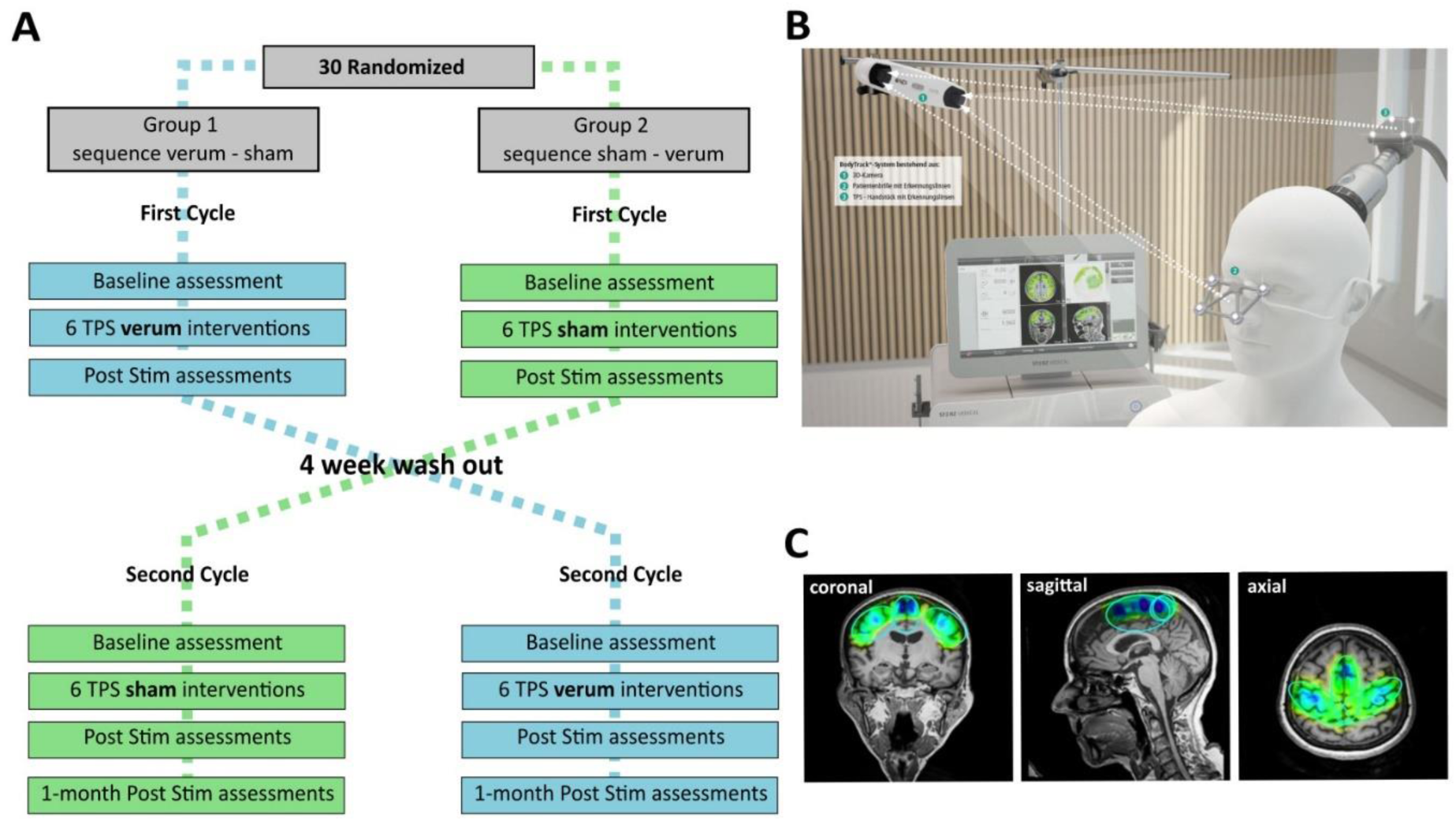
Study design. **(A)** In this randomized sham-controlled crossover investigation, 30 patients with Parkinson’s disease were randomized to the treatment sequence groups Verum-Sham and Sham-Verum. All participants received six verum and six sham transcranial pulse stimulation (TPS) sessions, along with comprehensive clinical assessments at baseline, post stimulation, and one month following the stimulations. The one-month post stimulation assessment of the first treatment cycle served as baseline for the second cycle. **(B)** TPS setting consisting of an ultrasound pulse generator connected with a hand piece for transcranial applications. Precise targeting was achieved by infrared sensors locating the handpiece and the patients’ head in real-time. Reproduced with permission from Storz Medical AG. **(C)** TPS targets, comprising the primary motor and somatosensory cortex, and the supplementary motor area, were individually defined on the patients’ anatomical magnetic resonance images.

We hypothesized that verum TPS treatment would outperform sham in reducing MDS-UPDRS-III score and enhancing coin rotation performance, the co-primary outcomes of the trial. Based on prior imaging findings with TPS in healthy participants and patients with Alzheimer’s disease [16–18], we anticipated increased task-based brain activation and improved white matter integrity.

## Methods

### Study design

In this randomized, double-blind, sham-controlled, crossover clinical trail, 30 patients with PD were included. The study was conducted at the Medical University of Vienna and obtained approval of the local ethics committee (EK1227-2015) and relevant regulatory authorities. Following comprehensive eligibility screening, participants were randomly allocated to the treatment sequence groups Verum-Sham, receiving verum TPS in the first treatment cycle and sham TPS in the second cycle (n = 15), or Sham-Verum (n =15, Figure 1A). The randomization was conducted using a randomization formula in Excel by a member of the study organization team who was not involved in outcome assessments. Each treatment cycle started with baseline clinical, neuropsychological, and MRI assessments that were followed by six verum or six sham stimulation sessions on separate days, spanned over two weeks. Clinical post stimulation evaluations were performed in the following days (median = 1, range 0-10 days), and one month afterwards. The one-month post stimulation assessment of the first treatment cycle served as baseline for the second cycle, resulting in a washout-phase between both treatment conditions of four weeks. MRI measurements were conducted at baseline and one day after the last stimulation session (median = 1, range 0-8 days), leading to a total of four MRI sessions across both cycles.

### Participants

To be included in this trial, patients required a neurologist-confirmed diagnosis of PD with clinically stable fine skilled and/or gross motor deficits and had to be aged 18 years or older. Exclusion criteria were relevant intracerebral pathology unrelated to PD (e.g., brain tumor), hemophilia or other blood clotting disorders, thrombosis, ongoing anticoagulation therapy, cortisone treatment within six weeks prior the first stimulation session, MRI contraindications, non-compliance with the protocol, continued participation in other studies, pregnancy or breastfeeding. Participants continued their individual standard medication and were instructed to keep medication stable during their individual study participation. All participants signed written informed consent forms after the nature and possible consequences of the study were explained and received financial compensation.

### Transcranial pulse stimulation

In total, participants received six verum and six sham TPS interventions using the prototype of the NEUROLITH (modified DUOLITH SD1, Storz Medical AG, Tägerwilen, Switzerland, Figure 1B). This system has been evaluated regarding safety, feasibility, and efficacy in Alzheimer’s disease [16,18–23], PD [14,15], and other neuropsychiatric disorders [24–27]. TPS received the CE-mark approval for the treatment of Alzheimer’s disease in 2018 and Food and Drug Administration (FDA) approval for studies in the U.S.A. (Investigational Device Exemption). Per session, 6000 TPS pulses were delivered to motor network areas, including M1, primary somatosensory cortex (S1), and the supplementary motor area (SMA). These regions of interest (ROIs) were predefined ellipses of constant size that were adjusted to align with individual anatomy based on structural MRI scans (Figure 1C) by a neurologist and TPS expert (RB). Pulses were evenly distributed within the ROIs, ensuring proper targeting by real-time tracking using an infrared camera monitoring sensors on goggles and the TPS hand piece. The ultrashort TPS pulses (3 μs) with a multifrequency band predominantly between 0.05 and 0.45 MHz [17] were applied with a pulse repetition frequency of 4 Hz and 0.25-mJ/mm^2^ energy flux density (duty cycle = 0.000012). Adjusting for skull attenuation of 80-90% [16], typical maximum pulse characteristics were 24 mW/cm^2^ spatial peak temporal average intensity (I_SPTA_), 1.6 kW/cm^2^ spatial peak pulse average intensity (I_SPPA_), and ambient pressure–related positive (7.5 MPa) and negative (−6 MPa) peak pressures below the skull. Sham stimulation was achieved by blocking the ultrasound pulses with a cap on the TPS hand piece, which looked identical and produced a sound closely matching the verum condition. Experimenters delivering TPS were informed about sham or verum condition but were not involved in the evaluation of outcome parameters. The principal investigator, along with the clinical and neuropsychological evaluators, were blinded to group assignment. Immediately after each stimulation, patients were asked to rate perceived pain and pressure on the scalp using a visual analogue scale (0-10) and to estimate, if verum or sham was applied.

### Behavioral outcomes

The motor examination of MDS-UPDRS (part III, [28]) and the coin rotation task (Foki et al. 2016) were the primary outcome measures in this trial. The MDS-UPDRS-III score, calculated from 33 scores based on 18 items, assessed motor functions such as rigidity, resting and action tremor, bradykinesia, gait, and other motor impairment, rated from 0 (normal) to 4 (severe impairment) by movement disorder specialists (HZ, AS). While the MDS-UPDRS-III is the most widely used clinical assessment in PD trials, it may not fully capture subtle motor impairments in early stages or difficulties with fine motor skills [29]. To complement the MDS-UPDRS-III, manual dexterity was examined using a behavioral coin rotation task [30], in which the patients rotated a plastic 2 € coin as quickly as possible for 20 s. The task was performed alternately with the left and right hand, starting with two training trials per side prior to the experiment. If the patient dropped the coin during the task, the timer was paused. Per side, the highest count from four attempts was used as co-primary outcome variable.

Subjective ratings of motor aspects of daily living, provided by both the patient and someone close to them, were recorded using the MDS-UPDRS-II, with ratings from 0 (normal) to 4 (severe impairment). For both the self-assessment and the external ratings, the sum of the 13 MDS-UPDRS-II items was calculated and analyzed as secondary outcomes. Comprehensive clinical motor evaluations further included the Keyboard Test [31], the Nine-hole Peg Test [32], Timed up and go Test [33], Buttoning and Unbuttoning Test [30], as well as the Parkinson’s disease Quality of Life Questionnaire (PDQ-39, [34]); data are not presented here.

The Montreal Cognitive Assessment (MoCA) is a screening instrument for mild cognitive impairments and comprises items on attention, concentration, executive functions, memory, speech, visuo-constructive abilities, calculation, conceptual thinking, and orientation. The maximum score is 30; a score of 26 and above indicates normal cognitive functioning. The MoCA corrected total score is adjusted for age, sex, and years of formal education (www.mocatest.ch/test/). For the assessment of depressive symptoms, the Beck Depression

Inventory (BDI-II, [35]), an 21-item questionnaire for self-evaluation with score ranges from 0 (no depression) to 63 (severe depression), and the short Geriatric Depression Scale (GDS-15, [36]) ranging from 0 (no depression) to 15 (severe depression) were used. Difficulties in the activities of daily living were evaluated using the Bayer Activities of Daily Living Scale (B-ADL, [37]), which comprises 25 items scored from 1 (no difficulties) to 10 (constant difficulties). The overall B-ADL score was calculated as the arithmetic mean of all available items. The Leisure behavior questionnaire (LBQ, adapted from [38]) comprised 25 items assessing the frequency of activities for entertainment, active movement, social interactions, creative activities, and cultural or educational activities on a scale from 0 (never) to 6 (daily). The average score across all 25 items entered statistical analysis. B-ADL and LBQ were intended to be completed by both the patient and someone familiar with the patient.

Initial outcome analysis of behavioral outcomes was performed by a contract research organization, applying T-tests of score changes (data not presented here). Predefined complex data analysis was conducted using SPSS Statistics (IBM Corp., version 29.0) and employed a repeated measures ANOVA (rmANOVA) with the within-subject factors Condition (Verum / Sham) and Session (Baseline, post stimulation, one-month post stimulation), testing for the interaction of interest Condition*Session for all behavioral outcomes. In case of a significant Condition*Session interaction, post-hoc comparisons between the post stimulation follow-ups (post stimulation, one-month post stimulation) and the baseline were calculated and corrected for multiple comparisons using Bonferroni-Holm adjustment. Data analysis was performed for the Intention-to-Treat sample (n = 30) with missing data being imputed using last-observation-carried-forward. The alpha level for significance was set at 0.025 for the primary outcomes, MDS-UPDRS-III and coin rotation, and at 0.05 for secondary outcomes. Partial eta squared (ηp^2^) was used to measure effect size, with thresholds of 0.01 for a small effect, 0.06 for a medium effect, and 0.14 for a large effect. In case of substantial outliers or skew data distribution, outcome variables were logarithmically transformed (LN(X +1)), to meet the assumptions for the rmANOVA. This procedure was successfully applied for the BDI-II, and the MDS-UPDRS-II (self and external). Transformation did not solve skewness for GDS and B-ADL data, so these outcome variables were analyzed using the non-parametric Friedman test for paired samples. When the Friedman test yielded a significant result, post-hoc pairwise comparisons were conducted using the Wilcoxon Signed Rank test, with Bonferroni-Holm correction applied for multiple comparisons.

Ratings of pressure and pain immediately after each TPS intervention were averaged over the six sessions of each condition and statistically compared using the non-parametric Wilcoxon Signed Rank test.

### MRI acquisition

For both experimental cycles, MRI sessions were recorded the week before and the week after TPS interventions. MRI measurements were performed using a 3 T SIEMENS PRISMA MRI with a 64-channel head coil. A T1-weighted structural image was recorded using a MPRAGE sequence (TE/TR = 2.7/1800 ms, inversion time = 900 ms, flip angle = 9°, resolution = 1 mm isotropic). A T2-weighted fluid-attenuated inversion recovery (FLAIR) sequence (TE/TR= 100/10000 ms, inversion time = 2500 ms, flip angle = 160°) and a T2-weighted 2D-fast-low-angle shot (FLASH) sequence (TE/ TR = 19.9/690 ms, flip angle = 20°) were applied to detect potential lesions, edemas or bleedings. DTI data were acquired using a whole-brain 30-direction gradient-echo-planar imaging (EPI) sequence (TE/TR = 95/10500 ms, multiband acceleration factor = 2, resolution = 2 mm isotropic, b-value = 1000 s/mm^2^) in anterior to posterior phase direction. For functional images, a T2*-weighted EPI sequence was used, with 64 slices aligned to the anterior-posterior commissure (AC-PC), covering the whole brain including cerebellum (TE/TR = 35/1400 ms, flip angle = 65°, multiband acceleration factor = 2, field of view = 224 × 224 mm, voxel size = 2 × 2× 2 mm, 10 % gap). For each of the functional runs, 125 volumes (2 min 55 s) were recorded and for resting state fMRI 430 Volumes (10 min 2 s, data not presented here).

### Functional brain activation

The fMRI motor task was adapted from previous work probing basal motor skills like finger tapping and more complex sensorimotor functions using a coin rotation paradigm for PD [39]. These tasks were used in an event-related design to differentiate motor planning, preparation, and execution. Each fMRI run consisted of four coin rotation and four finger tapping periods, performed with the right hand, with a fixed time for planning (2.8 s) and preparation (2.8 s), followed by motor execution phases (2.8 s, one finger tap or one 180° flip of a 2 € plastic coin) that were repeated 2-4 times. The order of the tasks was pseudo-randomized and time between the tasks varied between 5.6 to 8.4 s. The task and the motor phases were indicated by visually presented icons. The motor task consisted of eight runs which lasted approximately 3 minutes each.

Data preprocessing, first and second level statistics of fMRI data were performed using SPM12 (Version 7771; https://www.fil.ion.ucl.ac.uk/spm/software/spm12). Data preprocessing comprised slice time correction, spatial realignment across all four fMRI sessions, coregistration of functional and structural images, structural segmentation, normalization to Montreal Neurological Institute (MNI) space, and smoothing (6 mm FWHM Gaussian kernel). Data quality and accuracy of preprocessing procedures were checked on visual inspection. For first-level statistical analysis, a general linear model was used with the task events convolved with the hemodynamic response function. Data was corrected for subject motion (six motion parameters derived from the realignment procedure) and physiological noise (first three eigenvariates in non-cortical tissue as nuisance regressors). For each subject and session, contrasts were calculated for the tasks coin rotation and finger tapping (all phases). For group analysis, a flexible factorial model with the within-subject factors Condition (verum, sham), Session (pre / post stimulation) and Subject was applied for both tasks, coin rotation and finger tapping, separately. To estimate baseline motor activation, the first fMRI session was analyzed at the group level using a one-sample T-test against zero, conduced separately for each task.

For the complex motor task (coin rotation), an ROI analysis was conducted by extracting positive mean T-values for M1, SMA, and S1, as defined by the Harvard-Oxford atlas. To account for a potential acoustic bias of TPS application, the primary auditory cortex (bilateral TE 1.0, 1.1, and 1.2 (Jülich histological atlas: https://www.fz-juelich.de/de/inm/inm-7/leistungen/tools/jubrain-anatomy-toolbox, [40]) was also subjected to a control ROI analysis. Mean T-values were extracted from gray matter only, by confining the atlas ROI to individual gray matter areas, as derived by the structural segmentation procedure. Mean T-values of each ROI entered an rmANOVA using SPSS Statistics (IBM Corp., version 29.0) with Condition and Session as within-subject factors and the interaction between Condition and Session. In case of severe violations of the assumptions of normality, the non-parametric Friedman test was applied to analyze differences between conditions and sessions.

### Diffusion tensor imaging

Four DTI indices were investigated to assess white matter changes on whole brain level. As a measure of directionality of water diffusion, fractional anisotropy (FA) provides insight in white matter integrity [50]. Axial diffusivity (AD) measures water diffusion along the principal axis or the underlying fiber orientation, reflecting axon number and caliper (Kumar et al. 2012), while radial diffusivity (RD) indicates diffusion perpendicular to the tract, which is sensitive to myelin changes (Song et al. 2005), but is also influenced by diameter and density of axons [50]. Mean diffusivity (MD) reflects the average diffusion without directionality [51]. Intact neuronal microstructures, such as axonal cell membranes and myelin sheaths, promote increased levels of FA and reduce diffusivity values (MD, AD, RD) (Bennet and Madden 2014, Tae et al. 2018).

For processing and statistical analysis of DTI data, FMRIB Software Library (FSL) 6.0 and its toolboxes were used, following the online User Guide by Analysis Group of the Wellcome Centre for Integrative Neuroimaging (https://fsl.fmrib.ox.ac.uk/fsl/fslwiki/FSL). The initial step involved brain extraction and mask creation from the DTI b0 images utilizing the Brain Extraction Tool (BET) [41] with a threshold of 0.1. Next, FMRIB’s Diffusion Toolbox [42] was applied to perform eddy correction to minimize influence of artifacts and patients’ motion distortion [43]. Further, smoothing was conducted with fslmaths using a 1-voxel box kernel to promote data reliability [44]. After preprocessing, diffusion tensors were fitted to the images using DTI-FIT and visual inspection of all resulting images was performed to ensure data quality.

For all DTI indices, preprocessed images were statistically analyzed using Tract-Based Spatial Statistics (TBSS) [45]. This FSL tool enables voxelwise analysis through preprocessing, co-registration, normalization, mean image creation, and statistical modeling using a general linear model (GLM). These steps, done for each session and condition, involved removing outliers, nonlinear registration to the MNI152 standard space (1×1×1 mm) using the FMRIB58_FA template and its skeleton. To examine sham-controlled TPS effects, baseline-corrected contrasts (post stimulation vs. baseline) of verum versus sham condition and vice versa were created using fslmaths. These contrasts were then analyzed using the randomise toolbox with the threshold-free cluster enhancement option [46] with a one-sample T-test against zero, performing 5000 permutations with family-wise error (FWE) correction at p < 0.05. Variance smoothing (sigma = 5 mm) was applied due to the small sample size.

## Results

### Baseline characteristics

Between December 2019 and August 2022, 43 patients with PD were recruited from the outpatient clinic for movement disorders at the Department of Neurology, Medical University of Vienna, or via advertisements (see CONSORT flow diagram, **Figure S1**). Following comprehensive screening for eligibility, 30 patients were randomized to the treatment sequence groups Verum-Sham (first treatment cycle verum TPS, second cycle sham TPS, n = 15, Figure 1A) or Sham-Verum (n = 15). Two patients discontinued their participation prematurely. Both experienced COVID-19 infections during the first treatment cycle (both receiving sham TPS), one withdrew after completing the post-stimulation assessment, while the other withdrew following the one-month post-stimulation follow-up. Although both initially paused their participation due to the infections, they ultimately declined to resume and complete the study. Missing values were imputed using the last-observation-carried-forward method. In six additional patients, adherence to the per-protocol study schedule was not possible. Two patients experienced a prolonged interval between baseline and post-stimulation assessment in the first cycle. In four others, appointments for the second cycle were rescheduled. For these cases, the baseline assessments for the second cycle did not align with the 1-month post stimulation of the first cycle, so new baseline assessments were conducted at the actual start of the second cycle.

The intention-to-treat sample (n = 30) comprised eight female and 22 male participants with a mean age of 66.87 (SD = 7.03) and 14.30 (SD = 3.11) years of formal education, on average (**Table 1**). All patients were diagnosed with mild PD, with 27 patients showing a stage of 2 (90%), two patients a stage of 1 (6.7%), and another patient (3.3%) a stage of 2.5 on the Hoehn and Yahr Scale. Mean time since diagnosis was 5.30 (SD = 3.35) years.

**Table 1.**
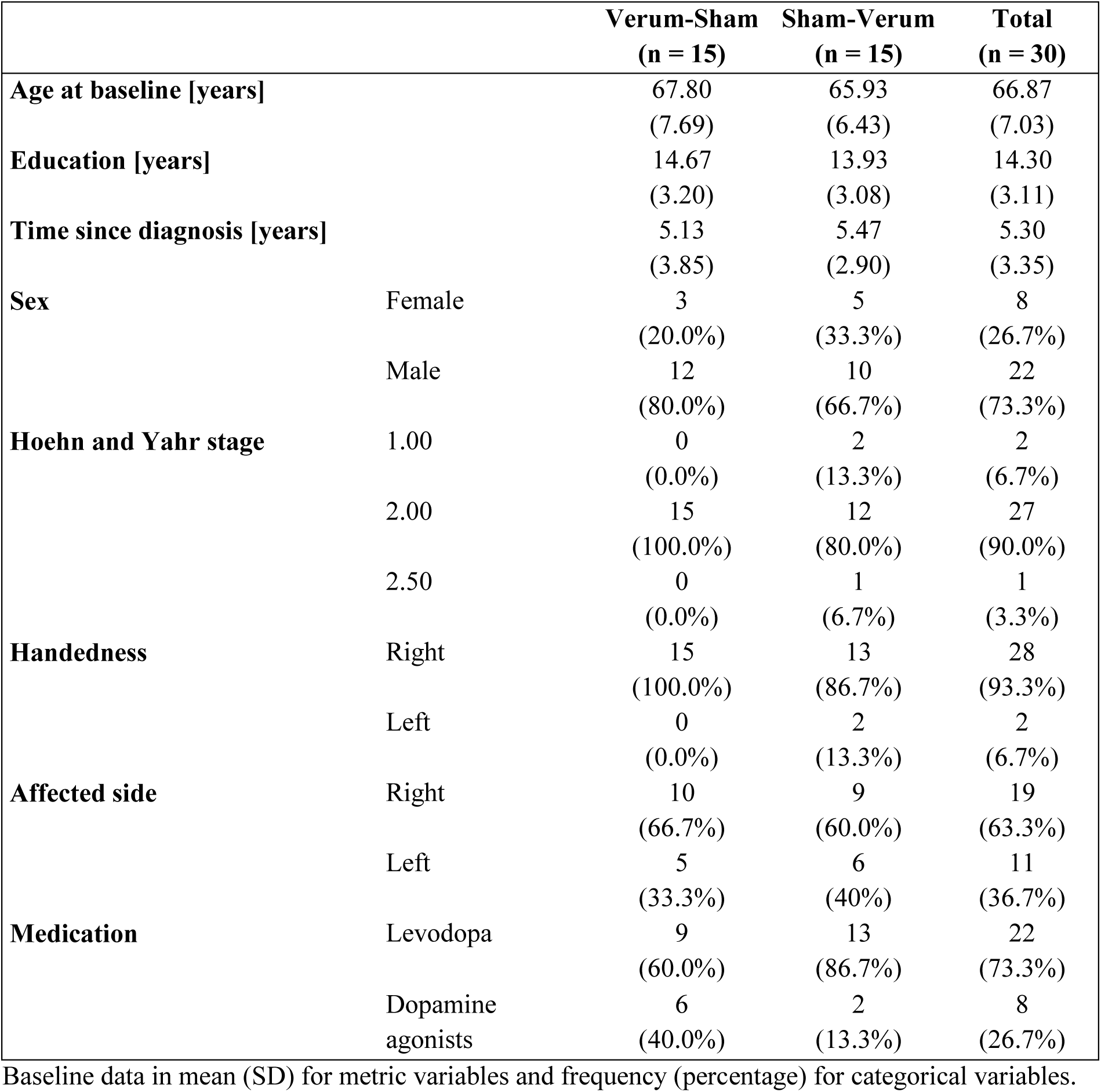
Baseline characteristics.

|  |  | **Verum-Sham**<br>**(n = 15)** | **Sham-Verum**<br>**(n = 15)** | **Total**<br>**(n = 30)** |
| --- | --- | --- | --- | --- |
| **Age at baseline [years]** |  | 67.80<br>(7.69) | 65.93<br>(6.43) | 66.87<br>(7.03) |
| **Education [years]** |  | 14.67<br>(3.20) | 13.93<br>(3.08) | 14.30<br>(3.11) |
| **Time since diagnosis [years]** |  | 5.13<br>(3.85) | 5.47<br>(2.90) | 5.30<br>(3.35) |
| **Sex** | Female | 3<br>(20.0%) | 5<br>(33.3%) | 8<br>(26.7%) |
|  | Male | 12<br>(80.0%) | 10<br>(66.7%) | 22<br>(73.3%) |
| **Hoehn and Yahr stage** | 1.00 | 0<br>(0.0%) | 2<br>(13.3%) | 2<br>(6.7%) |
|  | 2.00 | 15<br>(100.0%) | 12<br>(80.0%) | 27<br>(90.0%) |
|  | 2.50 | 0<br>(0.0%) | 1<br>(6.7%) | 1<br>(3.3%) |
| **Handedness** | Right | 15<br>(100.0%) | 13<br>(86.7%) | 28<br>(93.3%) |
|  | Left | 0<br>(0.0%) | 2<br>(13.3%) | 2<br>(6.7%) |
| **Affected side** | Right | 10<br>(66.7%) | 9<br>(60.0%) | 19<br>(63.3%) |
|  | Left | 5<br>(33.3%) | 6<br>(40%) | 11<br>(36.7%) |
| **Medication** | Levodopa | 9<br>(60.0%) | 13<br>(86.7%) | 22<br>(73.3%) |
|  | Dopamine agonists | 6<br>(40.0%) | 2<br>(13.3%) | 8<br>(26.7%) |

Baseline data in mean (SD) for metric variables and frequency (percentage) for categorical variables.

Twenty-two patients (73.3%) received levodopa, and eight (26.7%) took dopamine agonists (pramipexole, rotigotine, ropinirole) as standard medication. Most patients (93.3%) were right-handed and the right side was the more affected side in 19 patients (63.3%).

### Improved manual dexterity following verum TPS

Alongside the global motor impairment assessment using the MDS-UPDRS part III, (Goetz et al. 2008)), the coin rotation task [30] was a primary outcome of this trial. Interestingly, this assessment of manual dexterity illuminated a substantial difference between the right and the left hand (Figure 2). While the results for the right hand, being dominant in 93.3% of the patients, did not show significant results (compare **Table 2**), the interaction Condition*Session was significant for the left hand (p = 0.024, ηp^2^ = 0.14, large effect). Post-hoc tests for the interaction were significant for both follow-up sessions compared to baseline (p_corr_ = 0.048), confirming that the number of coin flips was significantly increased following verum compared to sham stimulation and this effect was maintained for at least one month.

**Figure 2.**
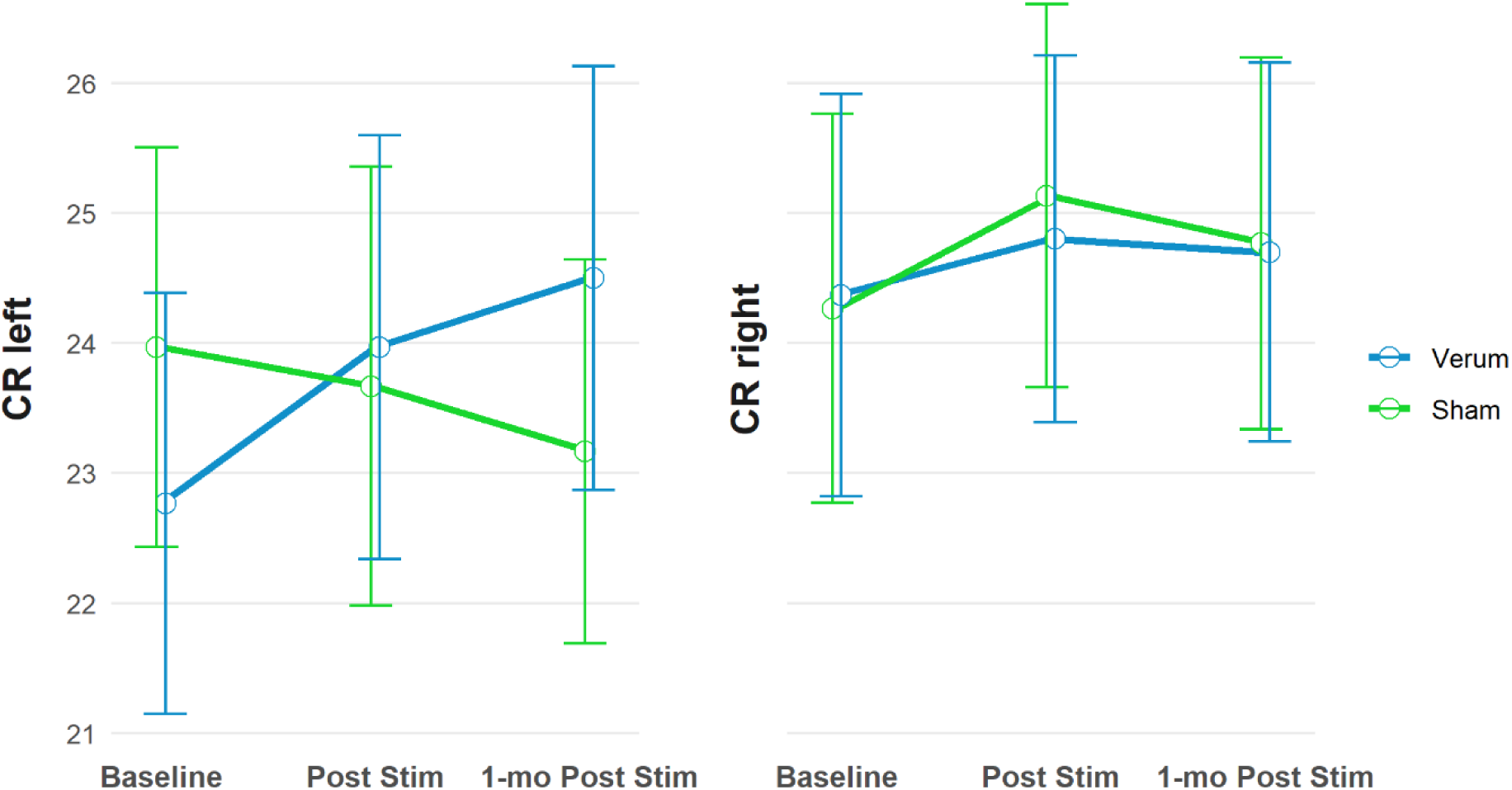
Coin rotation (CR) with the left and right hand. Performance for the left hand significantly improved following verum TPS treatment (Condition*Session interaction: p = 0.024, n = 30) indicating an improvement in manual dexterity lasting up to one month after the stimulation.

**Table 2.**
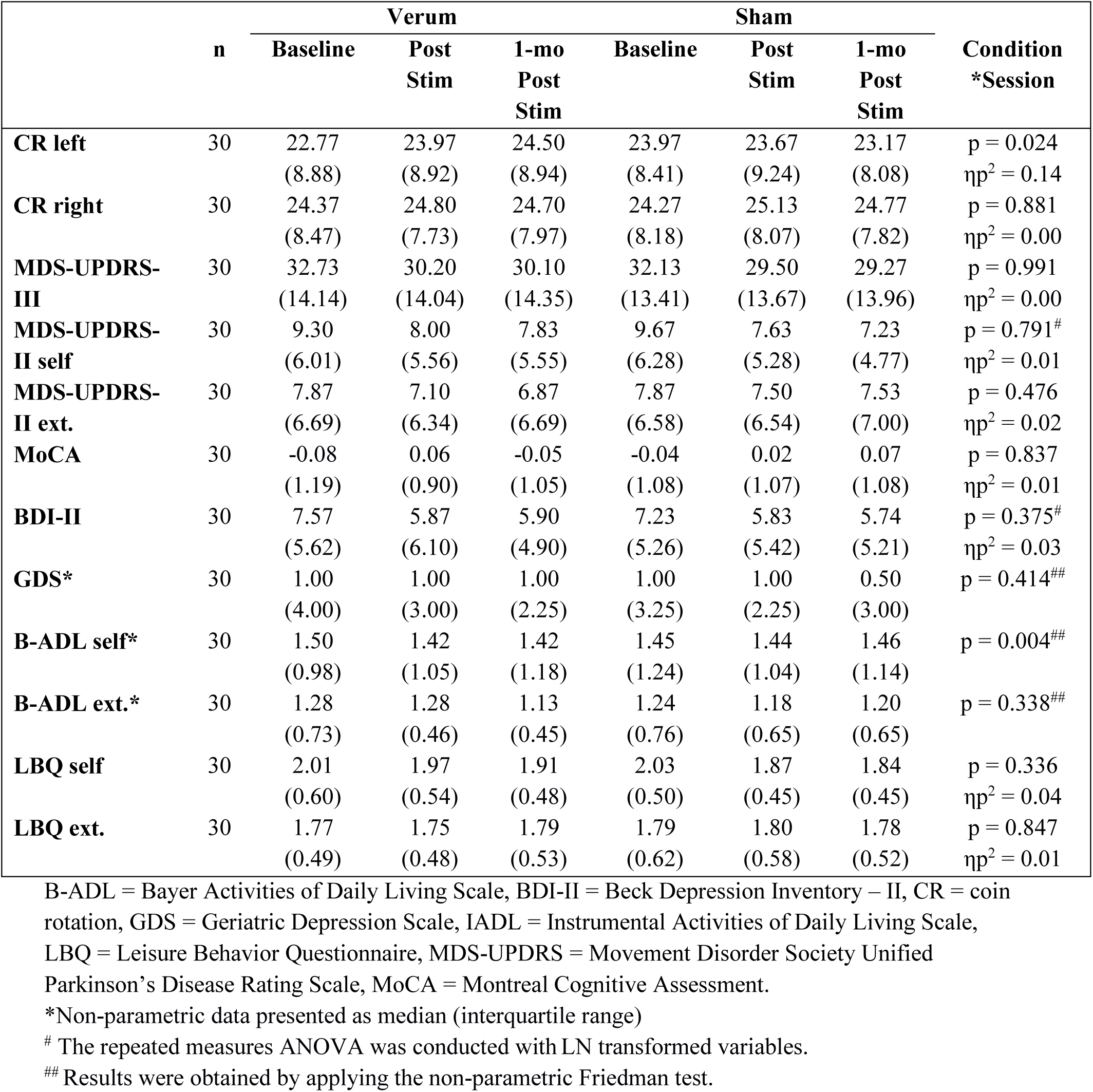
Outcome data as mean (standard deviation) at each time point and results of the interaction of interest (Condition*Session)

### No modulation of global motor scores by verum TPS

Overall, mean MDS-UPDRS-III scores decreased 4.93 points (SD = 9.23) from the first session (Cycle 1, baseline: mean = 34.10, SD = 13.38) to the last time point (Cycle 2, one-month post stimulation: mean = 29.17, SD = 14.43, Figure 3), indicating a clinically relevant score change in global motor symptoms over both cycles [47]. In the verum condition, mean MDS-UPDRS-III scores decreased from the baseline to one-month post stimulation by 2.63 points (32.73 (SD = 14.14) to 30.10 (SD = 14.35)) (**Table 2**). However, a comparable decrease of 2.86 points was found for the sham condition (baseline: 32.13 (SD = 13.41), one-month post stimulation: 29.27 (SD = 13.96)), resulting in a significant main effect of Session (p = 0.002, partial eta squared (ηp^2^) = 0.19, large effect), a non-significant main effect of Condition (p = 0.475, ηp^2^ = 0.02), and no significant interaction of interest (Condition*Session: p = 0.991, ηp^2^ = 0.00) in the rmANOVA.

**Figure 3.**
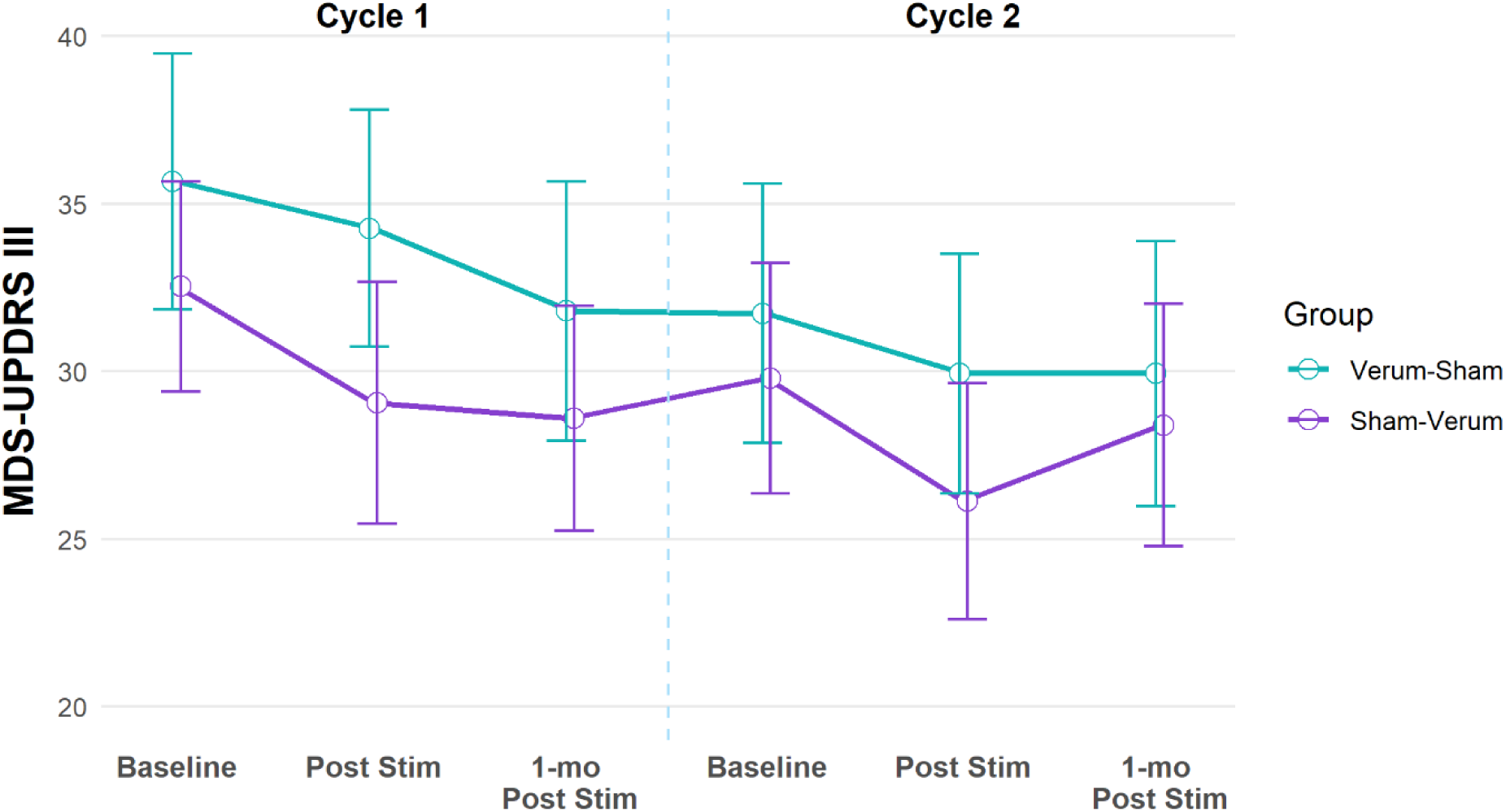
Change in the motor part of the Unified Parkinson’s disease rating scale (MDS-UPDRS-III) over time. The mean MDS-UPDRS-III scores decreased 4.93 points (SD = 9.23, n = 30) from the first session (Cycle 1, baseline) to the last time point (Cycle 2, one-month post stimulation), indicating a clinically relevant score change in global motor symptoms over time.

The MDS-UPDRS-II scores, as subjective estimation of motor impairments affecting activities of daily living, mirrored the results of the MDS-UPDRS-III. The self-estimation of motor impairments by the patients (MDS-UPDRS-II-self) decreased on average 3.67 points (SD = 3.51) during the study, starting with a mean score of 10.40 (SD = 6.65) in session 1 (Cycle 1, baseline) and ending with a mean score of 6.73 (SD = 4.91) in session 6 (Cycle 2, one-month post stimulation). The external assessment of the MDS-UPDRS-II, conducted by someone closely familiar with the patient, revealed a more subtle decrease from a mean score of 8.33 (SD = 6.43) in session 1 to 6.93 (SD = 6.87) in session 6. For both external and self-evaluation, the main effect of Session was significant but not the main effect of Condition or the interaction Condition*Session (see **Table 2** for details).

### Neuropsychological outcomes

The MoCA score assessing mild cognitive impairments remained stable throughout the study course and did not reveal any modulation by Condition or Session (**Table 2**). Depressive symptoms, assessed by the BDI-II, decreased on average 3.02 points (SD = 2.63) during the study participation, with mean scores of 8.67 (SD = 5.07) in session 1 and 5.64 (SD = 5.11) in session 6. The rmANOVA confirmed a significant effect of Session (p = 0.000, ηp^2^ = 0.40, large effect), but yielded no significance for Condition or Condition*Session (p = 0.375). Although the values for the GDS decreased descriptively as well, the non-parametric Friedman test failed to find a significant result. While the Friedman test analyzing external assessments of difficulties in activities of daily living (B-ADL external) was not significant, the patient ratings (B-ADL self) revealed a significant effect (p = 0.004). Post-hoc pairwise comparisons demonstrated a significant reduction in the B-ADL score after verum TPS (verum baseline: median = 1.50, IQR = 0.98, verum post stimulation: median = 1.42, IQR = 1.05, p_corr_ = 0.028), while this comparison was not significant for sham (sham baseline: median = 1.45, IQR = 1.24, sham post stimulation: mean = 1.44, IQR = 1.04, p_corr_ = 0.084).

The rmANOVA of the leisure behavior questionnaire showed a significant effect of Session with a decrease over both cycles in self-reported activities (LBQ-self), while the analysis of the external assessment (LBQ-external) did not reveal significant effects. For statistical details regarding neuropsychological outcomes, compare **Table 2**.

### Upregulation of functional motor activation following verum TPS

Functional brain activation during a complex dexterity task (coin rotation) and a basal motor task (finger tapping), both performed with the right hand, was subjected to a whole-brain analysis and an ROI analysis for all complete fMRI data sets (n =28).

As expected, the coin rotation task at baseline elicited activation in the left primary motor and somatosensory areas, contralateral to the hand moved, and in the bilateral anterior SMA. Small proportions of the right primary motor area were found to be activated as well, along with bilateral regions in superior and inferior frontal gyri (SFG, IFG), supramarginal gyri (SMG), occipital, and cerebellar regions. Deactivations were primarily located in regions of the default mode network (DMN) including the precuneus, lateral parietal and medial frontal areas, as well as bilateral temporal regions, IFG, insula, SFG and anterior cingulate cortex (ACC). Motor areas ipsilateral to the movement, such as the right posterior SMA, parts of right primary motor and somatosensory cortices, showed deactivations as well **(Figure S2).**

Subsequent result descriptions focus on the interaction of interest (Condition*Session), as changes to the respective baseline after verum and sham TPS are compared here. Since the motor task elicited not only widespread activation in the sensorimotor network, frontoparietal and visual areas, but also deactivation in well-known task-negative networks and beyond, the contrast of interest must be evaluated regarding changes relative to positive or negative T-values displayed in the first MRI session.

The interaction Condition*Session for the coin rotation task revealed an upregulation of somatosensory feedback processing, with activation found in the left postcentral gyrus and the left Rolandic operculum, for verum compared to sham TPS and increased activation in the motor network contralateral to the primarily engaged motor system (right M1, left cerebellum, Figure 4). In addition, a small cluster in the right ACC was found, a region engaged in motor coordination [48] and visuo-motor learning [49]. Negative clusters represent more pronounced deactivations in the verum compared to sham condition, particularly in the DMN (precuneus, angular gyrus). For more detail, see fMRI results table in the supplement (**Table S1).**

**Figure 4.**
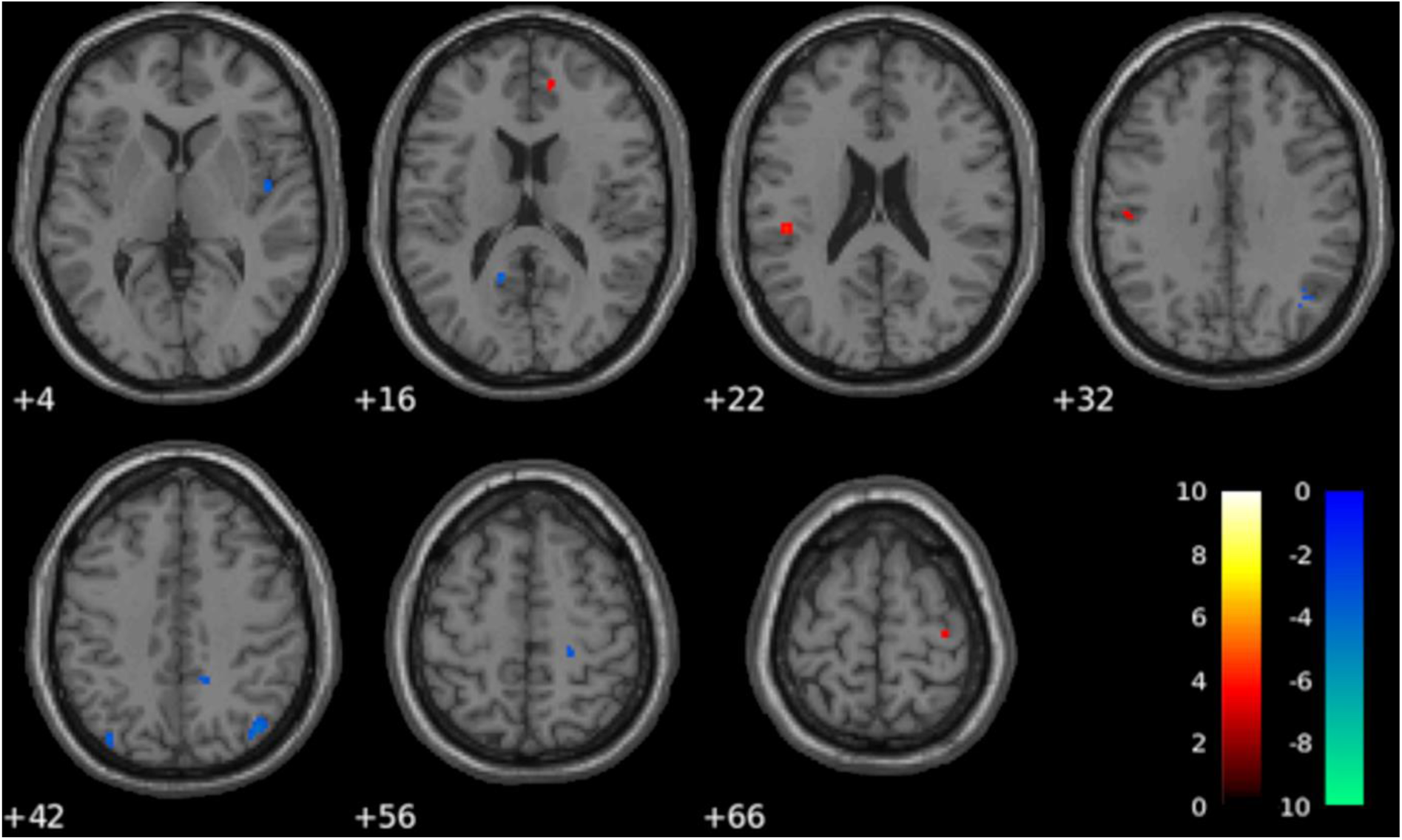
Change in functional motor activation in the coin rotation task. Increased functional activation in the primary and secondary somatosensory cortex, as well as in the right primary motor cortex (M1) and the anterior cingulate cortex, was observed following two weeks verum TPS treatment compared to sham in the coin rotation task (interaction Condition*Session, 0.001 uncorrected, k = 5, n = 28, verum > sham in red-to-yellow color scheme, sham > verum in green-to-blue).

At baseline, the right-hand finger tapping task elicited activation in the left primary motor and somatosensory cortices, as well as the bilateral SMA and IFG, right MFG, left insula, right SMG, along with bilateral visual and cerebellar areas. Deactivation was observed in regions of the DMN (precuneus, angular gyrus, medial frontal, temporal), in the ACC, superior frontal areas, right Rolandic operculum, occipital, and in small right precentral clusters (**Figure S3**).

For the finger tapping task, the interaction Condition*Session revealed several small clusters with increased activation verum vs. sham in the bilateral cerebellum, right parahippocampal gyrus, occipital lobe, superior frontal medial area, right precentral gyrus, and right middle cingulum but decreased activation in bilateral inferior parietal areas (**Table S2, Figure S4**). More pronounced deactivations were found in regions that showed subthreshold deactivation in the first MRI session (middle temporal, middle frontal, insula).

Functional brain activation was further analyzed in ROIs, comprising left and right M1, S1, and SMA. The rmANOVA assessing activation change in the coin rotation task revealed increased mean T-values in the verum condition across all investigated ROIs. A significant interaction of interest (Condition*Session) with a large effect size was observed for the in the right M1 (p = 0.005, ηp^2^ = 0.254) and the right S1 (p = 0.013, ηp^2^ = 0.209), see Figure 5 and **Table S3**. In addition, a significant main effect of Condition was observed in the right SMA, with higher mean T-values for verum compared to sham, and this difference was more pronounced in the post stimulation session. A control ROI analysis of potential coactivation of the primary auditory cortex showed no significant effects, neither in the coin rotation task (p = 0.440, Friedman-test), nor in the finger tapping task (p = 0.739, rmANOVA).

**Figure 5.**
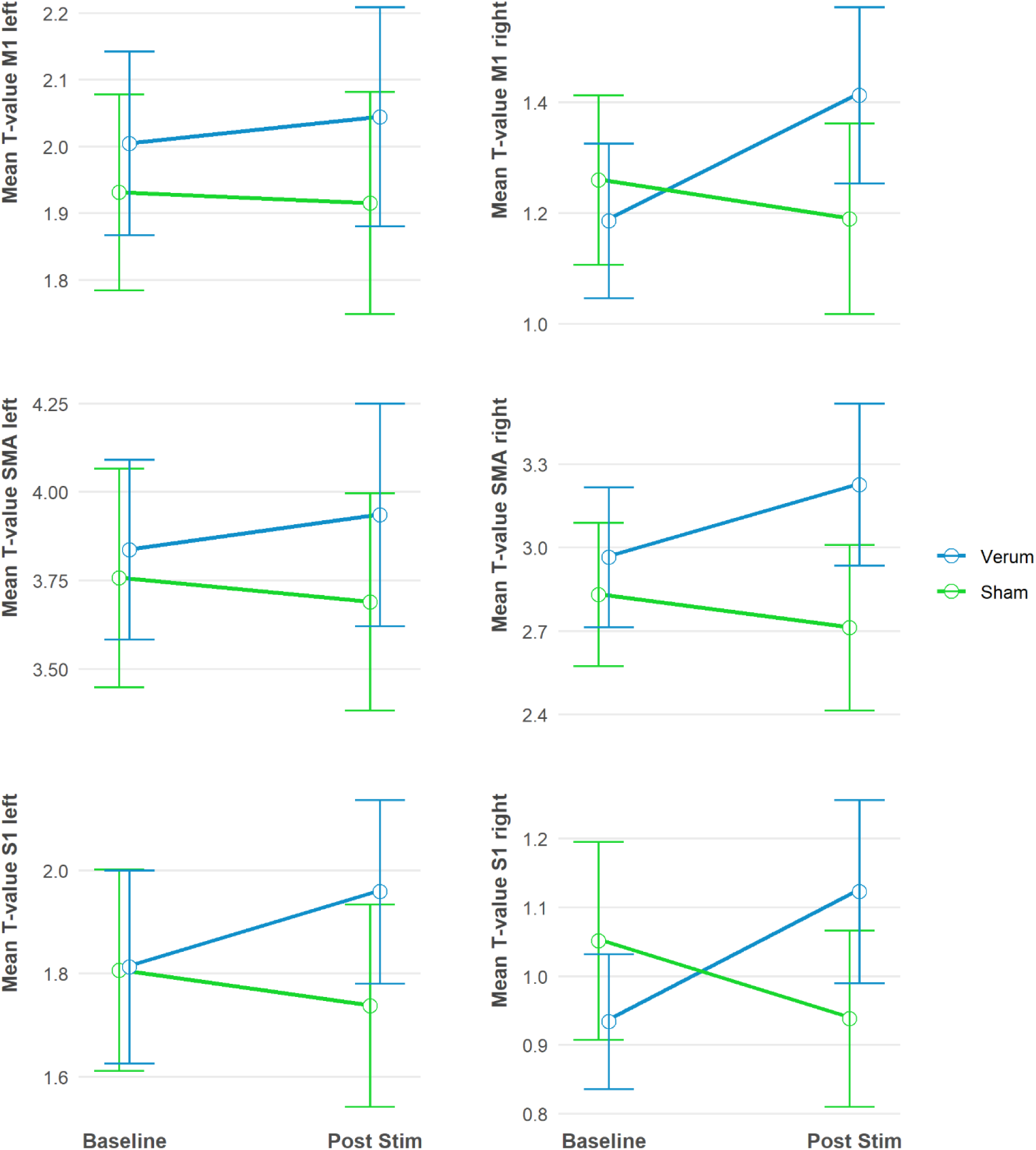
Functional activation during the coin rotation task in the sensorimotor regions of interest (ROIs). An upregulation of mean T-values following verum TPS (blue), compared to sham TPS (green), was observed across all ROIs. A significant Condition*Session interaction was found for the right primary motor cortex (M1, p =0.005, n = 28) and the right somatosensory cortex (S1, p = 0.013, n = 28), but not in the supplementary motor area (SMA).

### TPS enhanced structural integrity of sensorimotor white matter

Complete DTI data sets were available for 26 patients and analyzed using TBSS, revealing a significant increase in FA in the right postcentral gyrus following verum TPS compared to sham (26 voxel, peak coordinate: 29-33 61, p < 0.05 FWE corrected, Figure 6). For the other DTI indices, no significant change was noted (0.05 FWE corrected). Importantly, no area showed decreased FA or increased diffusivity (MD, AD, RD) following verum TPS.

**Figure 6.**
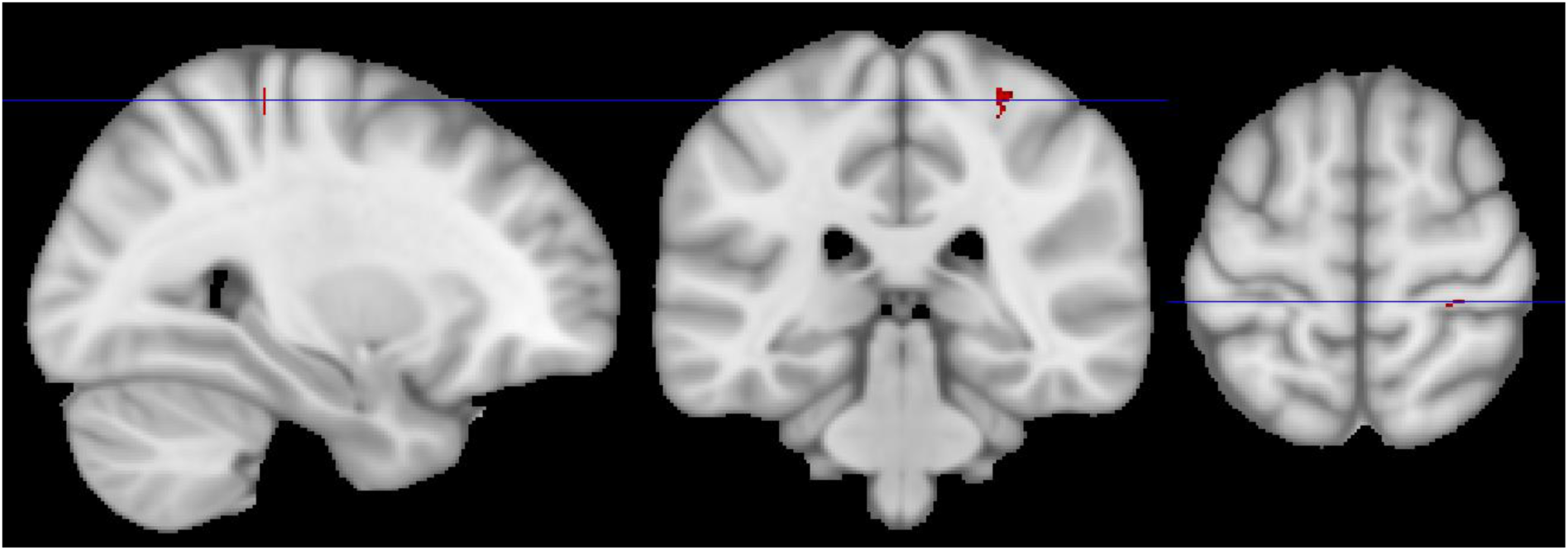
Diffusion tensor imaging. Increased fractional anisotropy (FA) was observed in right sensorimotor white matter tracts following two weeks of verum TPS treatment (verum > sham, post-stimulation > baseline, 0.05 FWE corrected, n = 26), indicating enhanced white matter integrity in stimulated areas.

### Adverse events

Ratings of side effects immediately after each stimulation demonstrated low levels of perceived pressure (verum: median = 0.83, IQR = 1.33; sham: median = 0.67, IQR = 1.00; p = 0.519, n = 28) and pain (verum: median = 0.00, IQR = 0.00; sham: median = 0.00, IQR = 0.00; p = 0.587, n = 28), without significant differences between verum and sham condition. Most of the stimulations were judged to be “real” (verum: 78.0%, sham: 78.6%), resulting in high sensitivity (78.0%) but low specificity (21.4%) and an accuracy of 49.7%, which is close to chance level in detecting the correct condition.

Overall, 21 adverse events (AEs) according to the Common Terminology Criteria for Adverse Events (CTCAE) were recorded during the study, although no serious adverse events occurred (see **Table S4**). Back, neck, hip, or Achilles’ tendon pain was reported by four patients (13.3%) at six occasions (2.5%), while muscle cramps were observed in three patients (10%, 1.7% of the time points). However, from a medical standpoint, a causal relationship to verum TPS appears unlikely. Depressive symptoms were noted in two patients (6.7%) at three time points (1.3%) and may be related to study procedures. Other symptoms potentially associated with TPS treatment were isolated cases of headache, tremor or fatigue (3.3% of the patients, 0.4% of the time points each).

Detailed neuroradiological evaluations, performed by an external radiologist (MS), revealed no neuropathological changes between baseline and post stimulation MR images.

## Discussion

In this randomized sham-controlled cross-over trial in 30 patients with PD, analysis of the primary outcome measures revealed a significant improvement in fine motor skills, as assessed by the coin rotation task, but no sham-controlled enhancement of gross motor abilities, evaluated by the MDS-UPDRS-III, following verum TPS.

Manual dexterity is the ability to use hands and fingers for complex and coordinated movements, including the manipulation of objects or tools, and is particularly important for activities of daily living [30]. Here, we found a specific improvement of fine motor skills, assessed by the coin rotation task, in the left hand following verum TPS. The right hand, being dominant in most patients, showed only limited response to TPS, potentially due to a ceiling effect for the skilled dominant hand. However, improved manual dexterity following verum TPS was reflected by significantly reduced self-reported difficulties in daily life, assessed by the B-ADL score.

Functional brain activation during the right-hand coin rotation task, requiring precise finger coordination in response to visual cues and somatosensory feedback, was assessed using fMRI at baseline and after verum and sham TPS. Contrasts of verum vs. sham TPS demonstrated significantly higher activation following verum TPS in the left S1 and Rolandic operculum, indicating an upregulation of primary and secondary somatosensory processing [52] in the competent left hemisphere (contralateral to the movement). Further, small clusters with higher activation following verum compared to sham stimulation were found in the ipsilateral M1 and the ACC. The ipsilateral M1 may support the dominant left M1 while rotating the coin with the right hand, as also reported for other complex manual tasks [53,54]. The ACC has been related to motor coordination and learning [48,49], as well as temporal discrimination of tactile stimuli [55]. Previous functional imaging findings in PD revealed that patients both *on* and *off* dopaminergic medication showed reduced S1 activation compared to controls during a coin rotation task, while M1 was upregulated by dopaminergic drugs [39]. On a behavioral level, coin rotation exhibited a poor dopaminergic response as well, while simple finger tapping movements, associated with bradykinesia, were improved by dopaminergic medication [56]. PD patients in this trial were on stable dopaminergic medication, receiving levodopa or dopaminergic agonists, which may have led to a saturation of the potential to further enhance motor functions. In contrast, primary and secondary somatosensory areas were upregulated by verum TPS suggesting a specific benefit to somatosensory processing, which typically shows limited responsiveness to dopaminergic drugs [39,56,57].

Significant findings regarding white matter integrity also focus on somatosensory areas, with significantly elevated FA levels in the right somatosensory gyrus following verum TPS. In general, a decrease in FA reflects reduced axonal alignment and demyelination, as observed in several neurodegenerative disorders, whereas an increase in FA is associated with improved white matter integrity and remyelination [50,58]. In the early stages of PD, FA is paradoxically increased in motor pathways, which has been attributed to compensatory reorganization in response to progressive nigrostriatal cell loss [59]. Specifically, PD patients in Hoehn & Yahr stage 1 display increased FA, compared to healthy controls and PD patients in Hoehn & Yahr stage 2, with lower FA values being correlated with motor severity in PD patients [60]. Compensatory axonal sprouting in nigrostriatal neurons was shown to be temporally associated with the reconstitution of normal motor behavior in animal models of PD and might explain the delayed onset of motor symptoms relative to the initial stages of cell loss and increased FA in initial disease stages [61]. The sham-controlled enhancement of FA in the stimulated sensorimotor system following verum TPS treatment in this study suggests that white matter integrity is promoted by TPS, potentially by inducing or reactivating compensatory neuroplasticity. An improvement of white matter integrity was also found in healthy participants following focal TPS of S1, with significantly reduced AD within the white matter tracts in the primary somatosensory and primary motor ROI [17]. While increased FA indicates improved overall fiber organization or integrity, decreased AD may reflect an increased axonal density or enlarged axonal calipers [62,63]. The differential TPS effects on FA and AD between PD patients and healthy participants may reflect PD-related neuropathology or age-related factors, given the older age of the PD cohort. However, since the stimulation protocols differed substantially between the two studies, direct comparisons are limited. Further research is needed to clarify the distinct effects of TPS across pathological and healthy states, as well as across age groups.

These behavioral, functional and structural imaging findings suggest that TPS is particularly effective at enhancing somatosensory processing in PD, which is essential for sensorimotor integration and coordination. Somatosensory dysfunction is a prominent feature of PD, even in its early stages, manifesting as deficits in temporal and spatial discrimination, spatial acuity, and tactile stimulus localization [64], all of which contribute to impaired dexterity and motor disturbances [65].

Nevertheless, motor symptoms such as rigidity, tremor, bradykinesia, and postural instability remain the cardinal symptoms of PD, and are traditionally assessed using Part III of the UPDRS or its revised version MDS-UPDRS [28]. The MDS-UPDRS-III decreased on average 4.93 points over the course of this study, meaning a clinically relevant benefit of overall study procedures [47]. With this 2% reduction in the MDS-UPDRS-III, the clinical improvement is comparable to findings in a retrospective clinical investigation in PD, reporting a decrease of 3.75 points (2%) in the UPDRS-III following TPS [14]. However, consistent with sham-controlled studies on the effects of tbFUS [11] and TPS [15] in PD, there was no significant difference between verum and sham TPS regarding this global motor score in the current study.

Although the MDS-UPDRS remains the most popular outcome measure in clinical trials in PD [66], several limitations must be addressed. First, the MDS-UPDRS-III is a noncontinuous and coarse rating scale with limitations regarding the detection of subtle motor difficulties, especially in early stages of the disease [29]. Second, within-subject reliability of the MDS-UPDRS-III is low, particularly in the *on* state, limiting its capability to track treatment efficiency or progression over time [67]. Third, fine motor skills are not adequately captured by the UPDRS-III [68] and its revision, leading to an underestimation of disabilities related to dexterity, particularly in early stages of PD. These limitations of the MDS-UPDRS may have contributed to the lack of a significant difference in this gross motor score between verum and sham TPS in the current study.

Another potential explanation may be the placebo effect, that is particularly pronounced in PD [69,70]. Placebo administration has been shown to increase the release of endogenous dopamine in the striatum in PD patients [69], with motor improvements specifically linked to dopamine release in the dorsal striatum [71]. While the expectation of clinical benefit activates the dopaminergic reward system in the ventral striatum, particularly the nucleus accumbens, translation into measurable motor improvement appears to require additional engagement of the dorsal striatum. Dopamine elevation in this region directly targets the core neuropathological substrate of PD: the depletion of dopamine within the nigrostriatal system [70,72]. The placebo response is influenced by the perceived invasiveness of an intervention, with high placebo rates in PD reported for surgical procedures of up to 55% [73]), 35-39% for DBS [74,75], and 17-46% for subcortical tissue ablation using high-intensity focused ultrasound [5,6]. Despite being noninvasive, brain stimulation techniques such as TMS or tDCS have been associated with prominent placebo responses in PD [76,77]. This may be attributed to the use of medical devices [78], the greater effort required as compared to drug administration, or a general response to the novelty of the intervention [73]. In addition, patients’ internal or externally induced positive treatment expectations have been shown to significantly influence treatment efficacy and the placebo response [75,76,79,80]. In the present study, PD patients demonstrated a strong belief in receiving verum TPS, with approximately 78% of the stimulation sessions rated as ‘real’, regardless of the actual condition. This high level of expectation likely contributed to a robust placebo response, potentially mediated by increased dopamine release in the striatum. Given that all participants were already receiving individually optimized dopaminergic therapy, additional dopamine release induced by positive expectation may have left limited room for further motor improvement through TPS itself.

To fully evaluate the potential of TPS in alleviating motor symptoms, it may be beneficial to study de novo PD patients who are not yet receiving dopaminergic treatment. Conversely, the intervention may also prove valuable in later stages of the disease, when dopaminergic therapies lose effectiveness and their side effects become more pronounced [81]. Moreover, non-motor symptoms, such as fatigue, cognitive impairments, and psychiatric disturbances, are more prevalent in advanced PD [82,83]. Targeting not only sensorimotor areas but also cognitive and affective networks with TPS may offer a more comprehensive therapeutic approach to address these complex symptoms, as previously demonstrated in a randomized controlled trial in Alzheimer’s disease [18].

Previous studies on focused ultrasound neuromodulation in PD have reported increased motor cortex excitability lasting up to 30 minutes following tbFUS of the primary motor cortex [12], as well as increased power in local field potentials in the basal ganglia for up to 40 minutes following FUS at 5 Hz (tbFUS) and 10 Hz [13]. Extending these short-term electrophysiological findings, we observed a sham-controlled upregulation of functional activation of the sensorimotor network, accompanied by enhanced white matter integrity in somatosensory regions, measured one day after six verum TPS sessions. These results suggest that repeated TPS of the sensorimotor cortex induces long-term neuroplasticity in the targeted network in PD.

Several limitations must be considered when interpreting the results of this study. Although the cross-over design, as applied here, has the advantage of a direct within-subject comparison between verum and sham stimulation, it bears the risk of carry-over effects. We administered a washout-phase of four weeks between both experimental cycles but still, we observed lower MDS-UPDRS baseline levels for the second cycle (**see** Figure 3), arguing for a carry-over effect. As both treatment sequence groups do not substantially differ in baseline characteristics and were similarly affected by the carry-over effect, overall results are not necessarily compromised. However, decreased baseline levels in the second cycle might have limited the full potential of verum TPS in the sham-verum group. Moreover, the repeated administration of the motor evaluations, cognitive tests, and questionnaires might have led to training or attrition effects.

As outlined in previous sections, the placebo effect is a particular concern in studies involving patients with PD. Therefore, it is crucial to employ a sham condition that closely matches the verum stimulation in appearance, acoustic characteristics, and sensory feedback. In our study, acoustic properties emitted by the TPS handpiece were roughly identical between the verum and sham stimulation, with both producing a repetitive ticking sound at 5 Hz, probably inducing habituation to this constant auditory input over the 20-minute stimulation period. Given the recent concerns about auditory biases in online assessments of MEPs [84], we conducted a control ROI analysis focusing on activation in the auditory cortex during the fMRI motor task. For both the complex coin rotation task and the simpler finger tapping task, mean T-values within the auditory ROI did not show significant results, indicating that differential recruitment of the auditory cortex is unlikely to account for the observed modulation of sensorimotor areas by verum TPS.

In addition, potential somatosensory confounds must be considered, as different sensations during verum or sham stimulation could have informed patients about the actual condition. However, there were no significant differences in the patients’ ratings of perceived pressure or pain on the scalp immediately following the verum and sham applications. Furthermore, patients’ ability to correctly identify the stimulation condition did not exceed chance level.

Regarding disease severity, we investigated a homogeneous sample, with 90% of the patients being classified as Hohn and Yahr stage 2. Consequently, generalizability of our findings to other disease stages is limited, as the underlying mechanisms of action of TPS may differ depending on disease progression. In earlier stages, TPS may primarily support compensatory neuroplasticity [60], whereas in later stages, add-on effects might become more prominent as the efficacy of dopaminergic therapy declines [81]. In addition, the influence of different PD subtypes and age of onset [85] was not considered in this investigation.

Prospective studies should address these limitations by including patients across different stages and subtypes of PD in randomized sham-controlled trials, ideally employing a parallel-group design with adequate sample size [86]. Comprehensive assessments of both gross and fine motor symptoms, as well as evaluations of somatosensory abilities, will be essential, preferably monitored over extended periods. The impact of dopaminergic therapy should also be systematically examined, for example by studying patients both *on* and *off* levodopa. Electrophysiological measurements, functional and structural imaging will help to elucidate the mechanisms of action of TPS on brain structure and function in PD. Finally, optimal TPS settings regarding stimulation parameter, target sites, and dosage protocols remain to be established to maximize treatment efficiency. With further refinement, TPS could serve as an additional therapeutic option to complement standard dopaminergic medications, aiming to enhance sensorimotor function and support activities of daily living in patients with PD.

## Data Availability

Access to deidentified participant data and study documents might be granted following review by the corresponding author and the Data Clearing House of the Medical University of Vienna.

## Acknowledgments

Regarding assistance in data acquisition, we thank Ahmad Amini, Benjamin Appel, Olga Ciobanu-Caraus, Anicca Egger, Fabian Ferdigg, Rosa Gredler, Daria Grigoryeva, Tabea Gruber, Deniz Inan, Patricia Jungwirth, Thomas Herpel, Lisa Kaindl, Nejla Karahasanović, Teodora Kolarova, Caroline Kolmanz, Francesca Lingenhel, Tudor Popescu, Patrik Russmann, Benjamin Spurny-Dworak, Anastasia Stashuk, and Saskia Tenk. We are grateful to Gregor Kasprian and Maria Schmook for the comprehensive neuroradiological evaluation.

## Funding

This work was supported by the Medical University of Vienna (grant SO10300020, to RB), the Herzfelder Foundation (to RB), and Storz Medical AG (to RB).

## Conflicts of interest

EM is a board member (auditor, unpaid) for the Austrian Society for functional MRI (ÖGfMRT). MM is a board member (secretary, unpaid) for the Austrian Society for functional MRI (ÖGfMRT). GD is a recipient of a DOC Fellowship of the Austrian Academy of Sciences at the Department of Psychiatry and Psychotherapy, Medical University of Vienna. RB received research and laboratory support from the Medical University of Vienna, the Austrian Science Fund, STORZ Medical AG, and the Herzfelder Foundation (Austria), and is president of scientific societies (unpaid). The other authors declare no competing interests.

## Notes

### Competing Interest Statement

EM is a board member (auditor, unpaid) for the Austrian Society for functional MRI (OeGfMRT). MM is a board member (secretary, unpaid) for the Austrian Society for functional MRI (OeGfMRT). GD is a recipient of a DOC Fellowship of the Austrian Academy of Sciences at the Department of Psychiatry and Psychotherapy, Medical University of Vienna. RB received research and laboratory support from the Medical University of Vienna, the Austrian Science Fund, STORZ Medical AG, and the Herzfelder Foundation (Austria), and is president of scientific societies (unpaid). The other authors declare no competing interests.

### Clinical Trial

NCT04333511

### Author Declarations

The Ethics committee of the Medical University of Vienna gave ethical approval for this work.

## References

[1] Postuma RB, Berg D, Stern M, Poewe W, Olanow CW, Oertel W, et al. MDS clinical diagnostic criteria for Parkinson’s disease. Mov Disord 2015;30:1591–601. 10.1002/mds.26424.

[2] Dickson DW, Braak H, Duda JE, Duyckaerts C, Gasser T, Halliday GM, et al. Neuropathological assessment of Parkinson’s disease: refining the diagnostic criteria. Lancet Neurol 2009;8:1150–7. 10.1016/S1474-4422(09)70238-8.

[3] Tanner CM, Ostrem JL. Parkinson’s Disease. N Engl J Med 2024;391:442–52. s10.1056/NEJMra2401857.

[4] Madrid J, Benninger DH. Non-invasive brain stimulation for Parkinson’s disease: Clinical evidence, latest concepts and future goals: A systematic review. J Neurosci Methods 2021;347:108957. 10.1016/j.jneumeth.2020.108957.

[5] Krishna V, Fishman PS, Eisenberg HM, Kaplitt M, Baltuch G, Chang JW, et al. Trial of Globus Pallidus Focused Ultrasound Ablation in Parkinson’s Disease. N Engl J Med 2023;388:683–93. 10.1056/NEJMoa2202721.

[6] Martínez-Fernández R, Máñez-Miró JU, Rodríguez-Rojas R, Del Álamo M, Shah BB, Hernández-Fernández F, et al. Randomized Trial of Focused Ultrasound Subthalamotomy for Parkinson’s Disease. N Engl J Med 2020;383:2501–13. 10.1056/NEJMoa2016311.

[7] Di Pino G, Pellegrino G, Assenza G, Capone F, Ferreri F, Formica D, et al. Modulation of brain plasticity in stroke: a novel model for neurorehabilitation. Nat Rev Neurol 2014;10:597–608. 10.1038/nrneurol.2014.162.

[8] Hu C, Zhang L, Luo G, Yao H, Song X, Liu Z. Clinical efficacy of low-intensity pulsed ultrasound in Parkinson’s disease with cognitive impairment. J Neurophysiol 2024;132:1633–8. 10.1152/jn.00323.2024.

[9] Beisteiner R, Hallett M, Lozano AM. Ultrasound Neuromodulation as a New Brain Therapy. Adv Sci 2023;10:e2205634. 10.1002/advs.202205634.

[10] Legon W, Strohman A. Low-intensity focused ultrasound for human neuromodulation. Nat Rev Methods Prim 2024;4:91. 10.1038/s43586-024-00368-6.

[11] Samuel N, Ding MYR, Sarica C, Darmani G, Harmsen IE, Grippe T, et al. Accelerated Transcranial Ultrasound Neuromodulation in Parkinson’s Disease: A Pilot Study. Mov Disord 2023;38:2209–16. 10.1002/mds.29622.

[12] Grippe T, Shamli-Oghli Y, Darmani G, Nankoo J-F, Raies N, Sarica C, et al. Plasticity-Induced Effects of Theta Burst Transcranial Ultrasound Stimulation in Parkinson’s Disease. Mov Disord 2024. 10.1002/mds.29836.

[13] Darmani G, Ramezanpour H, Sarica C, Annirood R, Grippe T, Nankoo J-F, et al. Individualized non-invasive deep brain stimulation of the basal ganglia using transcranial ultrasound stimulation. Nat Commun 2025;16:2693. 10.1038/s41467-025-57883-7.

[14] Osou S, Radjenovic S, Bender L, Gaal M, Zettl A, Dörl G, et al. Novel ultrasound neuromodulation therapy with transcranial pulse stimulation (TPS) in Parkinson’s disease: a first retrospective analysis. J Neurol 2024;271:1462–8. 10.1007/s00415-023-12114-1.

[15] Manganotti P, Liccari M, Maria Isabella Lombardo T, Della Toffola J, Cenacchi V, Catalan M, et al. Effect of a single session of transcranial pulse stimulation (TPS) on resting tremor in patients with Parkinson’s disease. Brain Res 2025;1850:149405. 10.1016/j.brainres.2024.149405.

[16] Beisteiner R, Matt E, Fan C, Baldysiak H, Schönfeld M, Philippi Novak T, et al. Transcranial Pulse Stimulation with Ultrasound in Alzheimer’s Disease—A New Navigated Focal Brain Therapy. Adv Sci 2019;7:1902583. 10.1002/advs.201902583.

[17] Matt E, Kaindl L, Tenk S, Egger A, Kolarova T, Karahasanović N, et al. First evidence of long-term effects of transcranial pulse stimulation (TPS) on the human brain. J Transl Med 2022;20:26. 10.1186/s12967-021-03222-5.

[18] Matt E, Mitterwallner M, Radjenovic S, Grigoryeva D, Weber A, Stögmann E, et al. Ultrasound Neuromodulation With Transcranial Pulse Stimulation in Alzheimer Disease: A Randomized Clinical Trial. JAMA Netw Open 2025;8:e2459170. 10.1001/jamanetworkopen.2024.59170.

[19] Cont C, Stute N, Galli A, Schulte C, Logmin K, Trenado C, et al. Retrospective real-world pilot data on transcranial pulse stimulation in mild to severe Alzheimer’s patients. Front Neurol 2022;13:948204. 10.3389/fneur.2022.948204.

[20] Shinzato GT, Assone T, Sandler PC, Pacheco-Barrios K, Fregni F, Radanovic M, et al. Non-invasive sound wave brain stimulation with Transcranial Pulse Stimulation (TPS) improves neuropsychiatric symptoms in Alzheimer’s disease. Brain Stimul 2024;17:413–5. 10.1016/j.brs.2024.03.007.

[21] Wojtecki L, Cont C, Stute N, Galli A, Schulte C, Trenado C. Electrical brain networks before and after transcranial pulsed shockwave stimulation in Alzheimer’s patients. GeroScience 2025;47:953–64. 10.1007/s11357-024-01305-x.

[22] Radjenovic S, Bender L, Gaal M, Grigoryeva D, Mitterwallner M, Osou S, et al. A retrospective analysis of ultrasound neuromodulation therapy using transcranial pulse stimulation in 58 dementia patients. Psychol Med 2025;55:e70. 10.1017/S0033291725000406.

[23] Fong TKH, Cheung T, Ngan STJ, Tong K, Lui WYV, Chan WC, et al. Transcranial pulse stimulation in the treatment of mild neurocognitive disorders. Ann Clin Transl Neurol 2023;10:1885–90. 10.1002/acn3.51882.

[24] Lohse-Busch H, Reime U, Falland R. Symptomatic treatment of unresponsive wakefulness syndrome with transcranially focused extracorporeal shock waves. NeuroRehabilitation 2014;35:235–44. 10.3233/NRE-141115.

[25] Cheung T, Li TMH, Lam JYT, Fong KH, Chiu LY, Ho YS, et al. Effects of transcranial pulse stimulation on autism spectrum disorder: a double-blind, randomized, sham-controlled trial. Brain Commun 2023;5:fcad226. 10.1093/braincomms/fcad226.

[26] Cheung T, Li TMH, Ho YS, Kranz G, Fong KNK, Leung SF, et al. Effects of Transcranial Pulse Stimulation (TPS) on Adults with Symptoms of Depression—A Pilot Randomized Controlled Trial. Int J Environ Res Public Health 2023;20:2333. 10.3390/ijerph20032333.

[27] Cheung T, Yee BK, Chau B, Lam JYT, Fong KH, Lo H, et al. Efficacy and safety of transcranial pulse stimulation in young adolescents with attention-deficit/hyperactivity disorder: a pilot, randomized, double-blind, sham-controlled trial. Front Neurol 2024;15:1364270. 10.3389/fneur.2024.1364270.

[28] Goetz CG, Tilley BC, Shaftman SR, Stebbins GT, Fahn S, Martinez-Martin P, et al. Movement Disorder Society-sponsored revision of the Unified Parkinson’s Disease Rating Scale (MDS-UPDRS): scale presentation and clinimetric testing results. Mov Disord 2008;23:2129–70. 10.1002/mds.22340.

[29] Regnault A, Boroojerdi B, Meunier J, Bani M, Morel T, Cano S. Does the MDS-UPDRS provide the precision to assess progression in early Parkinson’s disease? Learnings from the Parkinson’s progression marker initiative cohort. J Neurol 2019;266:1927–36. 10.1007/s00415-019-09348-3.

[30] Foki T, Vanbellingen T, Lungu C, Pirker W, Bohlhalter S, Nyffeler T, et al. Limb-kinetic apraxia affects activities of daily living in Parkinson’s disease: a multi-center study. Eur J Neurol 2016;23:1301–7. 10.1111/ene.13021.

[31] Goetz CG, Stebbins GT, Wolff D, DeLeeuw W, Bronte-Stewart H, Elble R, et al. Testing objective measures of motor impairment in early Parkinson’s disease: Feasibility study of an at-home testing device. Mov Disord 2009;24:551–6. 10.1002/mds.22379.

[32] Kellor M, Frost J, Silberberg N, Iversen I, Cummings R. Hand strength and dexterity. Am J Occup Ther Off Publ Am Occup Ther Assoc 1971;25:77–83.

[33] Podsiadlo D, Richardson S. The timed “Up & Go”: a test of basic functional mobility for frail elderly persons. J Am Geriatr Soc 1991;39:142–8. 10.1111/j.1532-5415.1991.tb01616.x.

[34] Berger K, Broll S, Winkelmann J, Heberlein I, Müller T, Ries V. Untersuchung zur Reliabilität der deutschen Version des PDQ-39: Ein krankheitsspezifischer Fragebogen zur Erfassung der Lebensqualität von Parkinson-Patienten. Aktuelle Neurol 1999;26:180–4. 10.1055/s-2007-1017628.

[35] Beck AT, Steer RA, Brown GK. Manual for the Beck Depression Inventory-II. San Antonio, TX: Psychological Corporation; 1996.

[36] Sheikh J, Yesavage J. Geriatric Depression Scale (GDS) Recent evidence and development of a shorter version. In: Brink T, editor. Clin. Gerontol. A Guid. to Assess. Interv., New York: The Haworth Press; 1986, p. 165–73.

[37] Hindmarch I, Lehfeld H, de Jongh P, Erzigkeit H. The Bayer Activities of Daily Living Scale (B-ADL). Dement Geriatr Cogn Disord 1998;9 Suppl 2:20–6. 10.1159/000051195.

[38] Hollneck U. Freizeitverhalten im höheren Lebensalter: Einflussfaktoren und Hemmnisse. 2009.

[39] Foki T, Pirker W, Geißler A, Haubenberger D, Hilbert M, Hoellinger I, et al. Finger dexterity deficits in Parkinson’s disease and somatosensory cortical dysfunction. Park Relat Disord 2015;21:259–65. 10.1016/j.parkreldis.2014.12.025.

[40] Morosan P, Rademacher J, Schleicher A, Amunts K, Schormann T, Zilles K. Human Primary Auditory Cortex: Cytoarchitectonic Subdivisions and Mapping into a Spatial Reference System. Neuroimage 2001;13:684–701. 10.1006/nimg.2000.0715.

[41] Smith SM. Fast robust automated brain extraction. Hum Brain Mapp 2002;17:143–55. 10.1002/hbm.10062.

[42] Behrens TEJ, Woolrich MW, Jenkinson M, Johansen-Berg H, Nunes RG, Clare S, et al. Characterization and propagation of uncertainty in diffusion-weighted MR imaging. Magn Reson Med 2003;50:1077–88. 10.1002/mrm.10609.

[43] Reese TG, Heid O, Weisskoff RM, Wedeen VJ. Reduction of eddy-current-induced distortion in diffusion MRI using a twice-refocused spin echo. Magn Reson Med 2003;49:177–82. 10.1002/mrm.10308.

[44] Madhyastha T, Mérillat S, Hirsiger S, Bezzola L, Liem F, Grabowski T, et al. Longitudinal reliability of tract-based spatial statistics in diffusion tensor imaging. Hum Brain Mapp 2014;35:4544–55. 10.1002/hbm.22493.

[45] Archer DB, Vaillancourt DE, Coombes SA. A Template and Probabilistic Atlas of the Human Sensorimotor Tracts using Diffusion MRI. Cereb Cortex 2018;28:1685–99. 10.1093/cercor/bhx066.

[46] Winkler AM, Ridgway GR, Webster MA, Smith SM, Nichols TE. Permutation inference for the general linear model. Neuroimage 2014;92:381–97. 10.1016/j.neuroimage.2014.01.060.

[47] Sánchez-Ferro Á, Matarazzo M, Martínez-Martín P, Martínez-Ávila JC, Gómez de la Cámara A, Giancardo L, et al. Minimal Clinically Important Difference for UPDRS-III in Daily Practice. Mov Disord Clin Pract 2018;5:448–50. 10.1002/mdc3.12632.

[48] Wenderoth N, Debaere F, Sunaert S, Swinnen SP. The role of anterior cingulate cortex and precuneus in the coordination of motor behaviour. Eur J Neurosci 2005;22:235–46. 10.1111/j.1460-9568.2005.04176.x.

[49] Engel A, Bangert M, Horbank D, Hijmans BS, Wilkens K, Keller PE, et al. Learning piano melodies in visuo-motor or audio-motor training conditions and the neural correlates of their cross-modal transfer. Neuroimage 2012;63:966–78. 10.1016/j.neuroimage.2012.03.038.

[50] Tae WS, Ham BJ, Pyun SB, Kang SH, Kim BJ. Current Clinical Applications of Diffusion-Tensor Imaging in Neurological Disorders. J Clin Neurol 2018;14:129–40. 10.3988/jcn.2018.14.2.129.

[51] Bennett IJ, Madden DJ. Disconnected aging: Cerebral white matter integrity and age-related differences in cognition. Neuroscience 2014;276:187–205. 10.1016/j.neuroscience.2013.11.026.

[52] Eickhoff SB, Grefkes C, Zilles K, Fink GR. The somatotopic organization of cytoarchitectonic areas on the human parietal operculum. Cereb Cortex 2007;17:1800–11. 10.1093/cercor/bhl090.

[53] Chettouf S, Rueda-Delgado LM, de Vries R, Ritter P, Daffertshofer A. Are unimanual movements bilateral? Neurosci Biobehav Rev 2020;113:39–50. 10.1016/j.neubiorev.2020.03.002.

[54] Suzuki T, Higashi T, Takagi M, Sugawara K. Hemispheric asymmetry of ipsilateral motor cortex activation in motor skill learning. Neuroreport 2013;24:693–7. 10.1097/WNR.0b013e3283630158.

[55] Pastor MA, Day BL, Macaluso E, Friston KJ, Frackowiak RSJ. The functional neuroanatomy of temporal discrimination. J Neurosci Off J Soc Neurosci 2004;24:2585–91. 10.1523/JNEUROSCI.4210-03.2004.

[56] Gebhardt A, Vanbellingen T, Baronti F, Kersten B, Bohlhalter S. Poor dopaminergic response of impaired dexterity in Parkinson’s disease: Bradykinesia or limb kinetic apraxia? Mov Disord 2008;23:1701–6. 10.1002/mds.22199.

[57] Nelson AJ, Hoque T, Gunraj C, Chen R. Altered somatosensory processing in Parkinson’s disease and modulation by dopaminergic medications. Parkinsonism Relat Disord 2018;53:76–81. 10.1016/j.parkreldis.2018.05.002.

[58] Alexander AL, Lee JE, Lazar M, Field AS. Diffusion tensor imaging of the brain. Neurother J Am Soc Exp Neurother 2007;4:316–29. 10.1016/j.nurt.2007.05.011.

[59] Mole JP, Subramanian L, Bracht T, Morris H, Metzler-Baddeley C, Linden DEJ. Increased fractional anisotropy in the motor tracts of Parkinson’s disease suggests compensatory neuroplasticity or selective neurodegeneration. Eur Radiol 2016;26:3327–35. 10.1007/s00330-015-4178-1.

[60] Wen M-C, Heng HSE, Ng SYE, Tan LCS, Chan LL, Tan EK. White matter microstructural characteristics in newly diagnosed Parkinson’s disease: An unbiased whole-brain study. Sci Rep 2016;6:35601. 10.1038/srep35601.

[61] Arkadir D, Bergman H, Fahn S. Redundant dopaminergic activity may enable compensatory axonal sprouting in Parkinson disease. Neurology 2014;82:1093–8. 10.1212/WNL.0000000000000243.

[62] Kumar R, Nguyen HD, Macey PM, Woo MA, Harper RM. Regional brain axial and radial diffusivity changes during development. J Neurosci Res 2012;90:346–55. 10.1002/jnr.22757.

[63] Takahashi M, Ono J, Harada K, Maeda M, Hackney DB. Diffusional Anisotropy in Cranial Nerves with Maturation: Quantitative Evaluation with Diffusion MR Imaging in Rats. Radiology 2000;216:881–5. 10.1148/radiology.216.3.r00se41881.

[64] Conte A, Khan N, Defazio G, Rothwell JC, Berardelli A. Pathophysiology of somatosensory abnormalities in Parkinson disease. Nat Rev Neurol 2013;9:687–97. 10.1038/nrneurol.2013.224.

[65] Lee MS, Lyoo CH, Lee MJ, Sim J, Cho H, Choi YH. Impaired finger dexterity in patients with parkinson’s disease correlates with discriminative cutaneous sensory dysfunction. Mov Disord 2010;25:2531–5. 10.1002/mds.23304.

[66] Lo C, Arora S, Lawton M, Barber T, Quinnell T, Dennis GJ, et al. A composite clinical motor score as a comprehensive and sensitive outcome measure for Parkinson’s disease. J Neurol Neurosurg & Psychiatry 2022;93:617 LP – 624. 10.1136/jnnp-2021-327880.

[67] Evers LJW, Krijthe JH, Meinders MJ, Bloem BR, Heskes TM. Measuring Parkinson’s disease over time: The real-world within-subject reliability of the MDS-UPDRS. Mov Disord 2019;34:1480–7. 10.1002/mds.27790.

[68] Koop MM, Shivitz N, Brontë-Stewart H. Quantitative measures of fine motor, limb, and postural bradykinesia in very early stage, untreated Parkinson’s disease. Mov Disord 2008;23:1262–8. 10.1002/mds.22077.

[69] de la Fuente-Fernández R, Ruth TJ, Sossi V, Schulzer M, Calne DB, Stoessl AJ. Expectation and dopamine release: mechanism of the placebo effect in Parkinson’s disease. Science 2001;293:1164–6. 10.1126/science.1060937.

[70] Quattrone A, Barbagallo G, Cerasa A, Stoessl AJ. Neurobiology of placebo effect in Parkinson’s disease: What we have learned and where we are going. Mov Disord 2018;33:1213–27. 10.1002/mds.27438.

[71] de la Fuente-Fernández R, Phillips AG, Zamburlini M, Sossi V, Calne DB, Ruth TJ, et al. Dopamine release in human ventral striatum and expectation of reward. Behav Brain Res 2002;136:359–63. 10.1016/s0166-4328(02)00130-4.

[72] de la Fuente-Fernández R, Schulzer M, Stoessl AJ. Placebo mechanisms and reward circuitry: clues from Parkinson’s disease. Biol Psychiatry 2004;56:67–71. 10.1016/j.biopsych.2003.11.019.

[73] Goetz CG, Wuu J, McDermott MP, Adler CH, Fahn S, Freed CR, et al. Placebo response in Parkinson’s disease: comparisons among 11 trials covering medical and surgical interventions. Mov Disord 2008;23:690–9. 10.1002/mds.21894.

[74] De La Fuente-Fernández R. Uncovering the hidden placebo effect in deep-brain stimulation for Parkinson’s disease. Park Relat Disord 2004;10:125–7. 10.1016/j.parkreldis.2003.10.003.

[75] Mercado R, Constantoyannis C, Mandat T, Kumar A, Schulzer M, Stoessl AJ, et al. Expectation and the placebo effect in Parkinson’s disease patients with subthalamic nucleus deep brain stimulation. Mov Disord 2006;21:1457–61. 10.1002/mds.20935.

[76] Strafella AP, Ko JH, Monchi O. Therapeutic application of transcranial magnetic stimulation in Parkinson’s disease: the contribution of expectation. Neuroimage 2006;31:1666–72. 10.1016/j.neuroimage.2006.02.005.

[77] Okabe S, Ugawa Y, Kanazawa I. 0.2-Hz repetitive transcranial magnetic stimulation has no add-on effects as compared to a realistic sham stimulation in Parkinson’s disease. Mov Disord 2003;18:382–8. 10.1002/mds.10370.

[78] Kaptchuk TJ, Goldman P, Stone DA, Stason WB. Do medical devices have enhanced placebo effects? J Clin Epidemiol 2000;53:786–92. 10.1016/s0895-4356(00)00206-7.

[79] Mameli F, Zirone E, Girlando R, Scagliotti E, Rigamonti G, Aiello EN, et al. Role of expectations in clinical outcomes after deep brain stimulation in patients with Parkinson’s disease: a systematic review. J Neurol 2023;270:5274–87. 10.1007/s00415-023-11898-6.

[80] Pollo A, Torre E, Lopiano L, Rizzone M, Lanotte M, Cavanna A, et al. Expectation modulates the response to subthalamic nucleus stimulation in Parkinsonian patients. Neuroreport 2002;13:1383–6. 10.1097/00001756-200208070-00006.

[81] Carrarini C, Russo M, Dono F, Di Pietro M, Rispoli MG, Di Stefano V, et al. A Stage-Based Approach to Therapy in Parkinson’s Disease. Biomolecules 2019;9. 10.3390/biom9080388.

[82] Goldman JG, Postuma R. Premotor and nonmotor features of Parkinson’s disease. Curr Opin Neurol 2014;27:434–41. 10.1097/WCO.0000000000000112.

[83] Broeders M, de Bie RMA, Velseboer DC, Speelman JD, Muslimovic D, Schmand B. Evolution of mild cognitive impairment in Parkinson disease. Neurology 2013;81:346–52. 10.1212/WNL.0b013e31829c5c86.

[84] Kop BR, Shamli Oghli Y, Grippe TC, Nandi T, Lefkes J, Meijer SW, et al. Auditory confounds can drive online effects of transcranial ultrasonic stimulation in humans. Elife 2024;12. 10.7554/eLife.88762.

[85] van Rooden SM, Heiser WJ, Kok JN, Verbaan D, van Hilten JJ, Marinus J. The identification of Parkinson’s disease subtypes using cluster analysis: a systematic review. Mov Disord 2010;25:969–78. 10.1002/mds.23116.

[86] Matt E, Radjenovic S, Mitterwallner M, Beisteiner R. Current State of Clinical Ultrasound Neuromodulation. Front Neurosci 2024;18. 10.3389/fnins.2024.1420255.

