## Supplement for "Transcranial Pulse Stimulation Enhances Dexterity in Parkinson’s Disease: A Randomized Sham-Controlled Clinical Trial"

Eva Matt[1], Nina Plischek[1], Michael Mitterwallner[1], Sonja Radjenovic[1], Alexandra Weber[1], Alina Domitner[1], Gregor Dörl[1], Anna Zettl[1], Sarah Osou[1], Anna Leitgeb[1], Agnes Santer[2], Viktoria Winkler[2], Julia Mandelburger[2], Heidemarie Zach[2], Roland Beisteiner[1*]

[1]Functional Brain Diagnostics and Therapy, Department of Neurology, Medical University of Vienna, Vienna, Austria

[2]Department of Neurology, Medical University of Vienna, Vienna, Austria

### Supplementary results

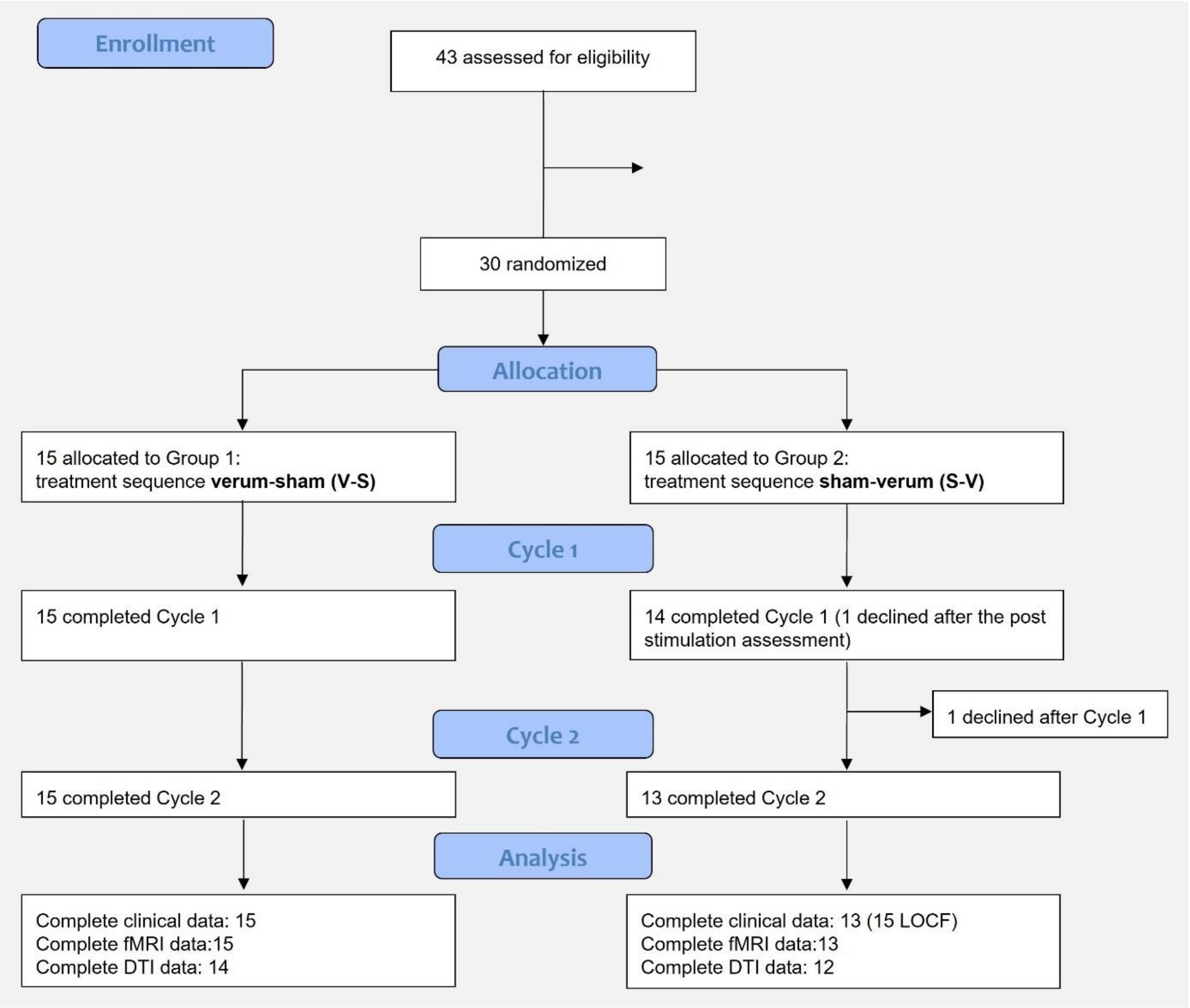


**Figure S1. CONSORT flow diagram.** Of the 43 patients with Parkinson's disease assessed for eligibility, 30 were randomized to the treatment sequence groups: Verum-Sham or Sham-Verum. Two patients discontinued the study prematurely, resulting in 28 complete data sets for clinical data, with missing values being imputed using the observation carried forward (LOCF) for the intention-to-treat analysis. Functional magnetic resonance imaging (fMRI) data were complete for 28 patients, and diffusion tensor imaging (DTI) data for 26 patients.

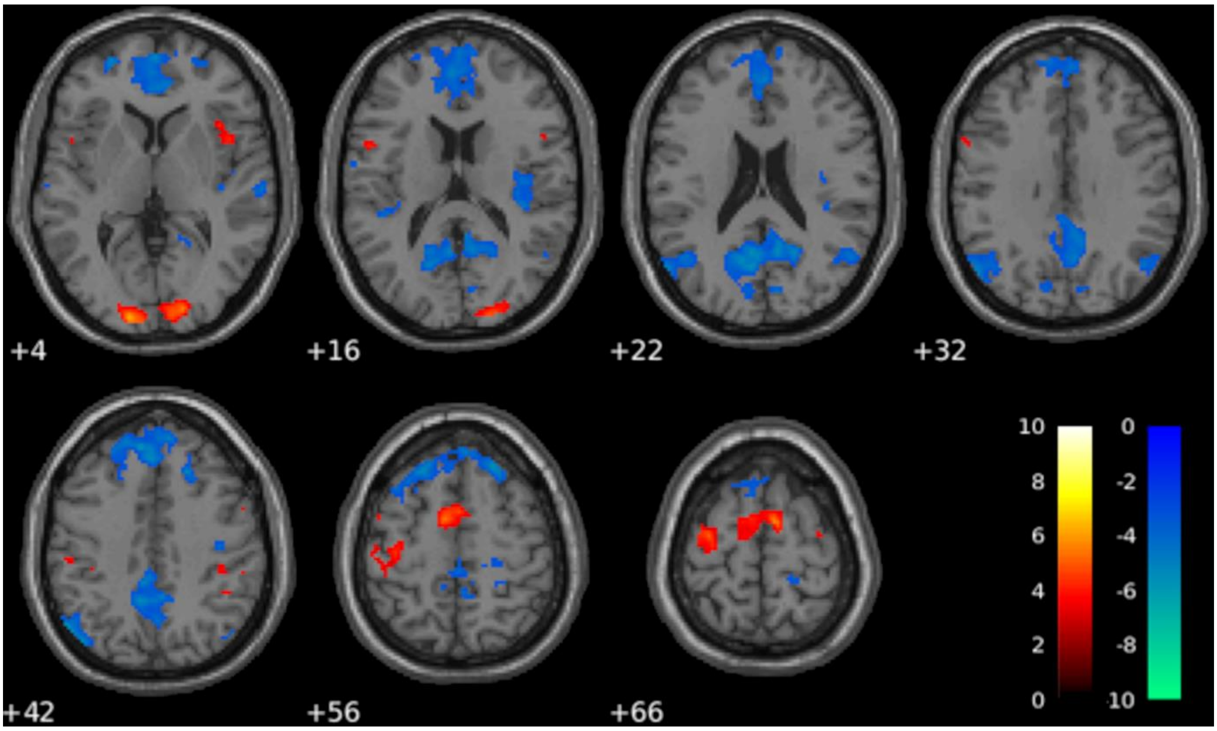


**Figure S2. Baseline coin rotation activation.** In the first functional magnetic resonance imaging (fMRI) session, coin rotation elicited activation in primary motor and somatosensory areas, as well as in the supplementary motor area, superior and inferior frontal gyri, supramarginal gyri (SMG), occipital, and cerebellar regions. Deactivations were predominantly found in regions associated with the default mode network (Session 1, T-test against 0, task-positive red-to-yellow, task-negative green-to-blue color scheme, 0.05 FWE corr., k = 5, n = 28).

**Table S1. Coin rotation task fMRI results with significant clusters for the interaction Condition*Session and the corresponding T-value in the first fMRI session**

| Interaction Condition*Session<br>Verum > Sham, Post Stim > Pre Stim<br>p = 0.001 uncorr., k = 5, n = 28 | | | | |
|---|---|---|---|---|
| **Area (AAL)** | **T** | **k** | **MNI-Coordinates** | **T-value in Session 1*** |
| MFG R | -3.6288 | 5 | 26 18 38 | -1.53 |
| ACC R | 3.7165 | 8 | 12 46 16 | -6.32 |
| MCC R | -4.3817 | 12 | 8 26 34 | -0.66 |
| PrecG (M1) R | 3.3751 | 5 | 34 -20 66 | 0.43 |
| PostcG (S1) L | 3.6987 | 9 | -50 -20 32 | 2.83 |
| PostcG (S1) R | -3.7002 | 7 | 22 -30 56 | -5.46 |
| RO L | 4.4058 | 26 | -46 -28 22 | -1.02 |
| Insula R | -3.8268 | 7 | 46 -6 4 | -0.42 |
| Precuneus L | -4.1891 | 13 | -14 -54 16 | -7.30 |
| | -3.8238 | 16 | -36 -76 40 | -5.88 |
| Precuneus R | -4.0849 | 17 | 14 -46 38 | -4.79 |
| AG R | -4.6548 | 137 | 44 -68 38 | -4.71 |
| Cerebellum L | 3.8502 | 13 | -12 -50 -18 | -1.37 |

* T-value displayed in the first MR session (T-test against 0) at the peak MNI coordinate of significant clusters found for the interaction Condition*Session.

ACC = Anterior cingulate cortex, AG = Angular gyrus, L = left, M1 = Primary motor cortex, MFG = Middle frontal gyrus, MCC = Middle cingulate cortex, PrecG = Precentral gyrus, PostcG = Postcentral gyrus, R = right, RO = Rolandic operculum, S1 = Primary somatosensory cortex.

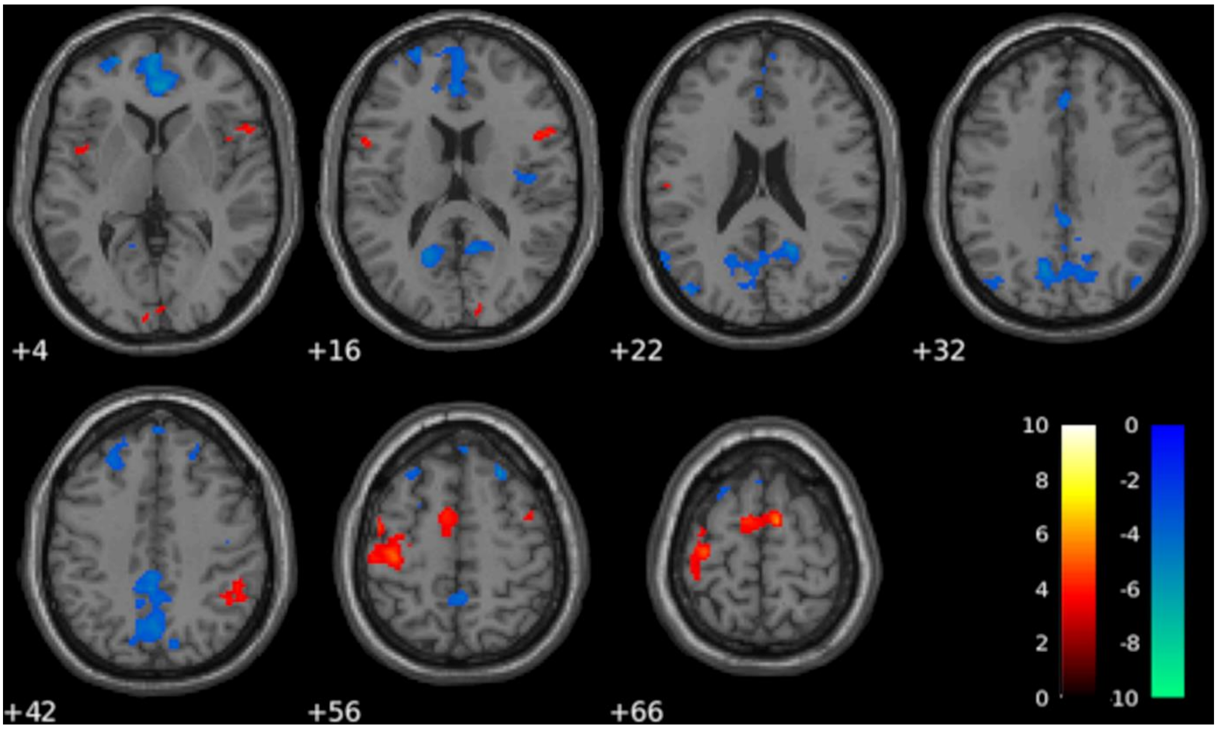


**Figure S3. Baseline finger tapping activation.** In the first fMRI session, primary motor and somatosensory cortex activation was observed during the right-hand finger tapping task, along with activation in the bilateral supplementary motor area, inferior and middle frontal gyri, insula, right supramarginal gyrus, visual and cerebellar areas and deactivations in the default mode network (Session 1, T-test against 0, task-positive red-to-yellow, task-negative green-to-blue color scheme, 0.05 FWE corr. k = 5, n = 28).

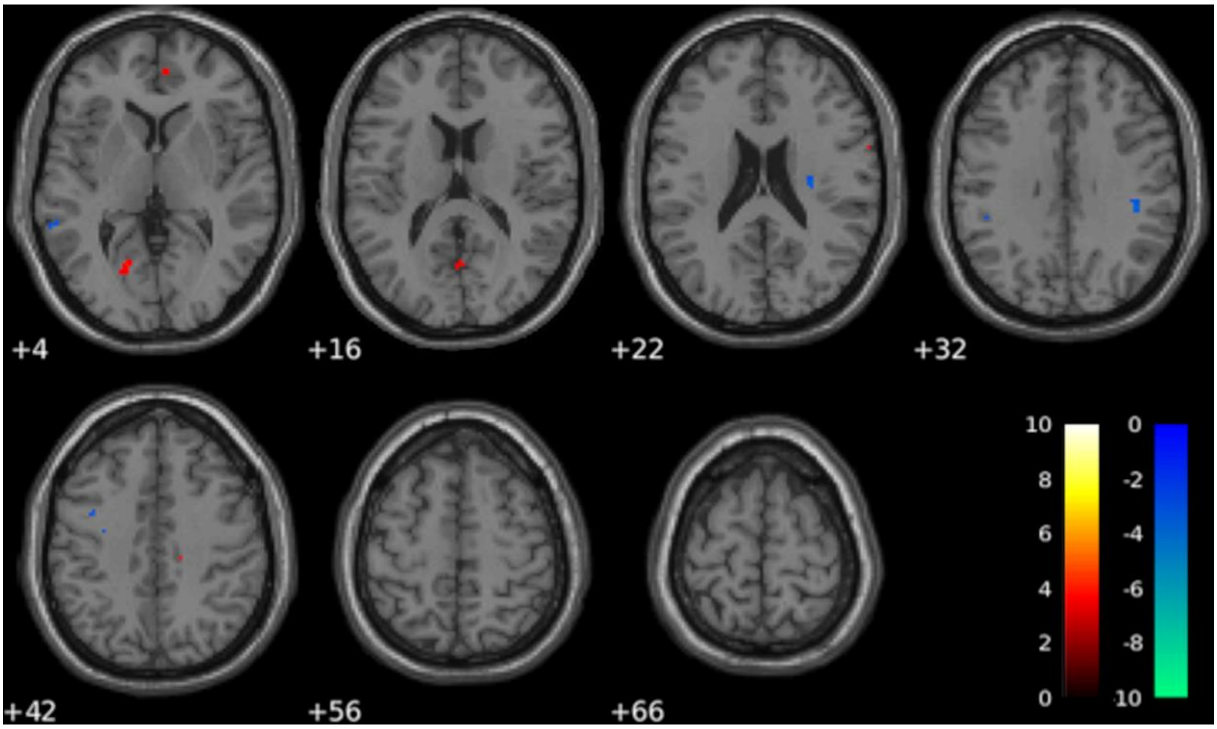


**Figure S4. Change in finger tapping motor activation.** Following verum TPS, increased activation in the right precentral gyrus, middle cingulum, superior frontal medial area, right parahippocampal gyrus, occipital lobe, and cerebellum, but decreased activation in bilateral inferior parietal areas was found (interaction Condition*Session, 0.001 uncorrected, k = 5, n = 28, verum > sham in red-to-yellow color scheme, sham > verum in green-to-blue).

**Table S2. Finger tapping task fMRI results with significant clusters for the interaction Condition*Session and the corresponding T-value in the first fMRI session**

| Interaction Condition*Session<br>Verum > Sham, Post Stim > Pre Stim<br>p = 0.001 uncorr., k = 5, n = 28 | | | | |
|---|---|---|---|---|
| **Area (AAL)** | **T** | **k** | **MNI-Coordinates** | **T-value in Session 1*** |
| SFM R | 3.4267 | 5 | 6 48 4 | -8.94 |
| MFG L | -4.6177 | 14 | -36 2 40 | -2.12 |
| MCC R | 3.6578 | 5 | 16 -24 42 | -0.81 |
| PrecG (M1) R | 3.6464 | 9 | 64 4 26 | 0.29 |
| Insula R | -3.7644 | 8 | 30 -18 22 | -0.68 |
| MTG L | -3.4597 | 7 | -60 -42 4 | -0.99 |
| LG L | 4.1031 | 35 | -16 -66 2 | -3.73 |
| SMG L | -3.4137 | 7 | -42 -40 36 | 2.32 |
| SMG R | -4.0931 | 17 | 42 -32 32 | 2.71 |
| ParahippG R | 3.5635 | 8 | 30 -26 -16 | -3.40 |
| IOG L | 3.3839 | 13 | -34 -84 -8 | -3.11 |
| Calcerine L | 4.2818 | 10 | -14 -64 12 | -6.41 |
| Calcerine R | 3.7214 | 10 | 2 -66 16 | -4.67 |
| Cerebellum L | 3.8148 | 12 | -26 -52 -22 | 3.95 |
| | 3.4208 | 5 | -22 -56 -18 | 3.47 |
| Cerebellum R | 3.9955 | 23 | 32 -56 -18 | 5.33 |

* T-value displayed in the first MR session (T-test against 0) at the peak MNI coordinate of significant clusters found for the interaction Condition*Session. IOG = Inferior occipital gyrus, L = left, LG = Lingual gyrus, M1 = Primary motor cortex, MFG = Middle frontal gyrus, MCC = Middle cingulate cortex, MTG = Middle temporal gyrus, ParahippG = Parahippocampal gyrus, PrecG = Precentral gyrus, R = right, SFM = Superior frontal medial, SMG = Supramarginal gyrus.

**Table S3. Mean T-value during the coin rotation task in the regions of interest**

| | Verum (n = 28) | | Sham (n = 28) | | Main effect of Condition | | Main effect of Session | | Interaction Condition* Session | |
|---|---|---|---|---|---|---|---|---|---|---|
| | **Baseline** | **Post Stim** | **Baseline** | **Post Stim** | **p** | **$\eta p^2$** | **p** | **$\eta p^2$** | **p** | **$\eta p^2$** |
| M1 left | 2.00 (0.73) | 2.04 (0.87) | 1.93 (0.78) | 1.92 (0.88) | 0.294 | 0.041 | 0.886 | 0.001 | 0.753 | 0.004 |
| M1 right | 1.19 (0.74) | 1.41 (0.84) | 1.26 (0.81) | 1.19 (0.91) | 0.324 | 0.036 | 0.161 | 0.071 | 0.005* | 0.254 |
| SMA left | 3.84 (1.34) | 3.93 (1.66) | 3.76 (1.63) | 3.69 (1.62) | 0.333 | 0.035 | 0.930 | 0.000 | 0.634 | 0.008 |
| SMA right | 2.97 (1.33) | 3.23 (1.54) | 2.83 (1.37) | 2.71 (1.57) | 0.035* | 0.154 | 0.645 | 0.008 | 0.168 | 0.069 |
| S1 left | 1.81 (0.99) | 1.96 (0.94) | 1.81 (1.03) | 1.74 (1.04) | 0.392 | 0.027 | 0.715 | 0.005 | 0.261 | 0.047 |
| S1 right | 0.93 (0.52) | 1.12 (0.70) | 1.05 (0.76) | 0.94 (0.68) | 0.760 | 0.004 | 0.606 | 0.010 | 0.013* | 0.209 |

Data is presented as mean and standard deviation in brackets. Main effects of Condition and Session and their interactions were tested with a repeated measures ANOVA, listing p-values and partial eta squared ($\eta p^2$). M1 = Primary motor cortex (precentral gyrus), S1 = Primary somatosensory cortex (postcentral gyrus), SMA = Supplementary motor area.

**Table S4. Adverse Events (AE) according to the Common Terminology Criteria for Adverse Events (CTCAE)**

| ID | Condition | Time point | AE (CTCAE term) | Grade | MedDRA Code | AE assessment |
|---|---|---|---|---|---|---|
| PD001 | Verum | Cycle 2, Baseline | Age-related vitreous detachment (Eye disorders - other specify) | | 10015919 | Causality not related (symptom occurred before the first verum stimulation), unexpected, transient |
| PD003 | Verum | Cycle 1, Post Stim | Back pain | | 10003988 | Causality not related, unexpected, transient |
| | Sham | Cycle 2, Post Stim | Flue like symptoms | 1 | 10016791 | Causality not related, unexpected, transient |
| PD015 | Verum | Cycle 2, Baseline | Depression | 2 | 10012378 | Causality with procedure possible (stress), causality with TPS not related (symptom occurred before the first verum stimulation), expected, transient |
| | Verum | Cycle 2, Post Stim | Candidiasis (Thrush) | 1 | 10043649 | Causality not related, unexpected, transient |
| | | **Cycle 2, Post Stim** | **Depression** | | **10012378** | **Causality possible, expected, transient** |
| PD016 | Verum | **Cycle 1, Post Stim** | **Headache** | **1** | **10019211** | **Causality possible, expected, transient** |
| | Sham | Cycle 2, Baseline | Back pain | | 10003988 | Causality not related, unexpected, transient |
| | Sham | Cycle 2, Post Stim | Intercostal neuralgia | | 10029223 | Causality not related, unexpected, transient |
| | | Cycle 2, 1-month Post Stim | Neck pain | | 10028836 | Causality not related, unexpected, transient |
| **PD018** | **Sham** | **Cycle 2, 1-month Post Stim** | **Depression** | **1** | **10012378** | **Causality possible, expected, transient** |
| PD019 | Sham | Cycle 2, Post Stim | Muscle cramp in the right thigh | | 10028294 | Causality not related, unexpected, transient |
| PD020 | Verum | Cycle 2, Post Stim | Hip pain (Bone pain) | | 10006002 | Causality not related, unexpected, transient |
| | **Sham** | **Cycle 2, Post Stim** | **Tremor in the right arm** | | **10044565** | **Causality possible, unexpected, transient** |
| PD028 | **Verum** | **Cycle 1, Post Stim** | **Fatigue** | | **10016256** | **Causality possible, unexpected, transient** |
| | Sham | Cycle 2, Post Stim | Sinusitis | | 10040753 | Causality not related, unexpected, transient |

| | | | | | |
|---|---|---|---|---|---|
| | Sham | Cycle 2, 1-month Post Stim | Muscle cramp in the legs | 10028294 | Causality not related, unexpected, transient |
| PD029 | Sham | Cycle 2, baseline | Muscle cramp in the legs | 10028294 | Causality not related (pre-existing), unexpected, transient |
| | Sham | Cycle 2, Post Stim | Muscle cramp | 10028294 | Causality not related (pre-existing), unexpected, transient |
| PD030 | Sham | Cycle 2, Post Stim | Pain in the Achilles' tendon | 10033371 | Causality not related, unexpected, transient |
| | Sham | Cycle 2, 1-month Post Stim | Pain in the Achilles' tendon | 10033371 | Causality not related, unexpected, transient |
